# Clozapine prescribing in Germany: temporal trends and regional variations, 2012–2022

**DOI:** 10.64898/2025.12.03.25341492

**Authors:** Oliver H. F. Scholle, Oliver Riedel, Mishal Qubad, Michael Dörks, Bianca Kollhorst, Robert A. Bittner, Christian J. Bachmann

## Abstract

Clozapine is the most effective and only approved drug in treatment-resistant schizophrenia (TRS). Studies based on data up to 2014 concluded that it is underutilized in most industrialized countries, including Germany. Since 2019, national guidelines explicitly recommend clozapine as first-line therapy in TRS. We aimed to assess whether clozapine use in Germany has increased in recent years and to examine regional variations. Using claims data covering about 20% of the German population (GePaRD), we calculated the yearly prescription prevalence and incidence of clozapine among individuals aged 0–64 years based on outpatient dispensations. For 2022, we also assessed regional variations in clozapine prescription prevalence at the district level (restricted to N=202 districts with ≥20,000 individuals). From 2012 to 2022, the overall (age- and sex-standardized) prescription prevalence of clozapine continuously decreased by 16% (from 77.6 to 65.5 per 100,000). The relative decline was greatest in women aged 30–39 years (−51%) and in men aged 30–34 years (−57%), in urban areas (large urban cities: -23%; urban districts: -16%), and in regions with high socioeconomic status (−22%). Over the same period, the overall prescription incidence of clozapine decreased by 41%. In 2022, regional clozapine prescription prevalence differed up to 39-fold. In conclusion, clozapine prescribing in Germany did not increase from 2012 to 2022, despite new clozapine-favoring guidelines, and showed substantial regional variation. Our results suggest a persisting underutilization of clozapine in most regions in Germany. Further research on barriers and facilitators on clozapine use in Germany is needed.

## INTRODUCTION

Clozapine remains the most effective antipsychotic for schizophrenia and the only medication specifically approved for patients with treatment-resistant schizophrenia (TRS) (Bittner et al., 2023). At least one-third of all individuals with schizophrenia fulfill TRS criteria (Diniz et al., 2023; Howes et al., 2017; Siskind et al., 2022). These patients bear a disproportionately high share of the disorder’s stigma, clinical burden, and associated healthcare costs (Kane et al., 2019), highlighting the need for adequate evidence-based treatment with clozapine.

Both, meta-analyses of randomized controlled trials and real-world cohort studies consistently demonstrate clozapine’s unique clinical benefits (Brodeur et al., 2022; Correll et al., 2022b; Huhn et al., 2019; Luykx et al., 2025; Masdrakis and Baldwin, 2023; Masuda et al., 2019; Qubad and Bittner, 2023; Taipale et al., 2020). Among all antipsychotics, clozapine shows the largest improvements in global psychopathology and in positive, negative, and depressive symptoms (Chakos et al., 2001; Kane et al., 1988; Mizuno et al., 2020; Wagner et al., 2021). Compared to other antipsychotics, clozapine elicits greater improvements in treatment adherence and lower rates of suicidal behavior (Brodeur et al., 2022; Hennen and Baldessarini, 2005; Huhn et al., 2019; Meltzer, 2012; Meltzer et al., 2003; Siris, 2001). Clozapine is also the most effective drug for reducing aggressive behavior, substance use, hospitalization rates, and for preventing further relapses in patients with a second psychotic episode (Brunette et al., 2006; Faay et al., 2018; Frogley et al., 2012; Lähteenvuo et al., 2022, 2021; Taipale et al., 2025).

Moreover, clozapine achieves greater reductions in all-cause and suicide mortality than any other antipsychotic in patients with schizophrenia (Correll et al., 2022b; Taipale et al., 2020; Wagner et al., 2021). Clozapine’s singular effect on all-cause mortality is partly attributable to its superior impact on treatment adherence for somatic comorbidities (Solmi et al., 2021). Further, clozapine has a lower risk of extrapyramidal side-effects (Huhn et al., 2019). Accordingly, current expert consensus guidelines recommend clozapine as a second-line treatment in cases of persistent positive symptoms accompanied by suicidality, aggression, extrapyramidal symptoms, or tardive dyskinesia (Wagner et al., 2020).

Crucially, a substantial proportion of TRS cases manifest during the first episode of psychosis (FEP), with approximately 25% of FEP patients meeting TRS criteria (Siskind et al., 2022). Delayed initiation of clozapine after the emergence of treatment-resistance—typically averaging 5–7 years—substantially lowers response rates (Jones et al., 2022; Shah et al., 2020; Yada et al., 2015). This underscores the critical importance of a timely detection of treatment resistance and immediate initiation of clozapine, already during the early stages of manifest illness.

However, contrary to national and international guideline recommendations (Correll et al., 2022a; Deutsche Gesellschaft für Psychiatrie und Psychotherapie, Psychosomatik und Nervenheilkunde, 2025; McCutcheon et al., 2025), clozapine remains considerably underutilized across most healthcare systems. Estimates from international studies suggest that in most countries, only a minority of eligible patients receive clozapine, with considerable variations both within and across countries (Bachmann et al., 2017; Bittner et al., 2023).

Among the available metrics to quantify drug utilization, the prescription prevalence (i.e., the number of persons with at least one prescription in a population) is essential. The most recent population-based study on trends in the prescription prevalence of clozapine, encompassing 17 countries including Germany, covered the period from 2005 to 2014 (Bachmann et al., 2017). Since then, long-term trend data on the prescription prevalence of clozapine have not been systematically investigated across Europe. Moreover, only few studies have examined fine-grained regional variation in clozapine prescribing.

Therefore, our aim was to comprehensively characterize longitudinal trends in outpatient clozapine prescribing in Germany from 2012 to 2022—both overall and stratified by age, sex, urbanicity, and area-level socioeconomic deprivation—and to quantify the extent of regional variations at the district level.

## METHODS

We conducted year-wise cross-sectional studies, analyzing routinely collected German healthcare data.

### Data source

We used claims data from four statutory health insurance providers in Germany (GePaRD), which include information on approximately 25 million persons who have been insured with one of the participating providers since 2004 or later (Haug and Schink, 2021). Per calendar year, the data cover approximately 20% of the general population representing all geographical regions of Germany. Available demographic data include sex, age, and district-level region of residence. Prescription data encompass all reimbursed medication prescribed in the outpatient setting by general practitioners or specialists. Prescriptions are coded according to the German modification of the WHO Anatomical Therapeutic Chemical (ATC) classification system (version as of April 2023 for this study).

### Study design

To be eligible for the year-wise study populations from 2012 to 2022, individuals had to meet the following criteria: (a) valid information on sex and an age between 0 and 64 years, (b) documented residency in Germany and (c) continuous insurance coverage throughout the respective calendar year allowing for coverage gaps of up to 30 days. Individuals who died or were born during the respective year were also included, provided they were continuously insured from January 1 until their date of death or from birth until December 31. For estimating the prescription incidence, eligible individuals additionally had to be continuously insured during the entire preceding calendar year and must not have received a clozapine prescription during that year. Persons aged 65 years or older were excluded to minimize the likelihood of capturing clozapine prescriptions for conditions such as psychosis related to Parkinson’s disease or various forms of dementia, which are more prevalent in older age groups (Bachmann et al., 2017).

### Clozapine prescriptions

For each year, we identified prescriptions of clozapine (ATC code N05AH02) based on reimbursed outpatient dispensations.

### Urbanicity and socioeconomic deprivation

District-level characteristics were linked to individuals via their region of residence (total number of districts: 401). Urbanicity was defined according to the official classification of district types based on settlement structure (as of 2017), categorized into four levels ranging from “large urban city” to “sparsely populated rural district”. This classification is based on settlement characteristics such as the proportion of the population in large and medium-sized cities, overall population density, and population density excluding large and medium-sized cities. Additionally, we analyzed trends by district-level socioeconomic deprivation using the publicly available German Index of Socioeconomic Deprivation (as of 2018) (Michalski et al., 2022).

### Data analysis

For each year from 2012 to 2022, we calculated the prescription prevalence of clozapine as the number of individuals with at least one prescription per 100,000 individuals. Using the same approach, we also calculated the prescription incidence, restricting the analysis to individuals with continuous insurance coverage (as previously defined) and no clozapine prescription in the preceding calendar year. All estimates were computed both overall and stratified by sex and age group. For prescription prevalence, additional stratifications were performed by urbanicity and district-level socioeconomic deprivation, the latter classified into quintiles; with the second to fourth quintile combined into a single intermediate category.

For the year 2022, we calculated the clozapine prescription prevalence at the district level. Due to small sample sizes, we limited the analysis to districts with a database population of at least 20,000 individuals, resulting in a total of 202 (out of 401) districts.

All prevalence and incidence estimates were calculated along with 95% confidence intervals (CIs) and were directly standardized by age and sex, using the German population of December 31, 2022 as the reference. All statistical analyses were performed using SAS version 9.4 (SAS Institute, Cary, NC, USA).

## RESULTS

From 2012 to 2022, the number of included individuals per year ranged from 11,653,312 (2012) to 13,671,786 (2022) (**Supplementary Table S1**).

### Temporal trends overall and by age and sex

In 2022, the overall age- and sex-standardized clozapine prescription prevalence was 65.5 per 100,000 (95% CI: 64.2; 66.9) (**Figure 1**; **Supplementary Table S1**). Prevalence was higher among males (79.2 per 100,000; 95% CI: 77.0; 81.3) than in females (51.5 per 100,000; 95% CI: 49.8; 53.2).

**Figure 1:**
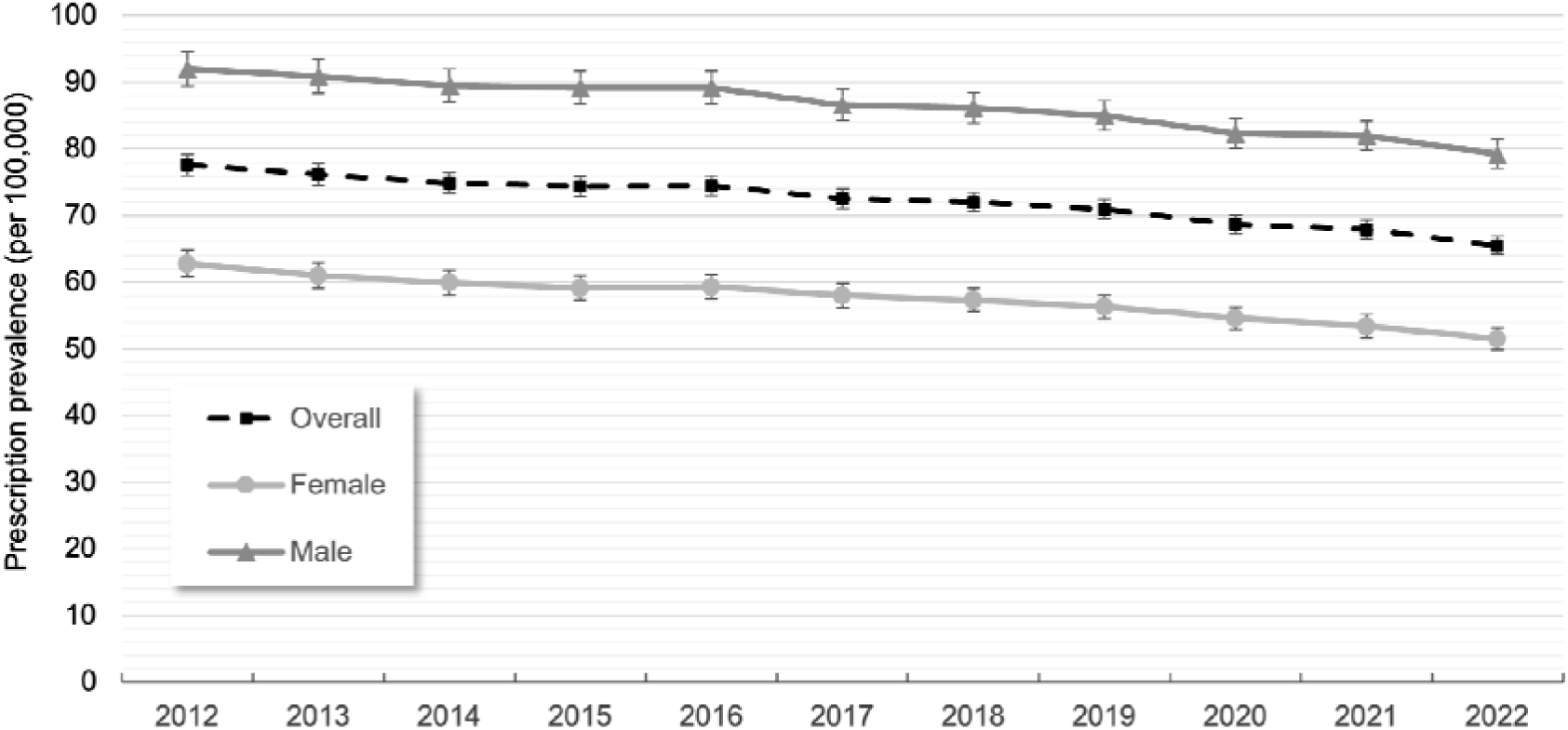
Standardized prescription prevalence (with 95% CIs) of clozapine overall and by sex between 2012 and 2022

Between 2012 and 2022, the overall age- and sex-standardized prescription prevalence of clozapine continuously decreased (−16%, from 77.6 to 65.5 per 100,000) (**Figure 1**). This downward trend was observed in both sexes, with an 18% decrease among females (from 62.8 to 51.5 per 100,000) and a 14% decrease among males (from 92.0 to 79.2 per 100,000). As shown in **Figure 2**, the age- and sex-standardized prescription incidence declined even more sharply than the prevalence. Between 2012 and 2022, the prescription incidence of clozapine decreased by 41%, from 7.1 to 4.2 per 100,000 (**Supplementary Table S2**).

**Figure 2:**
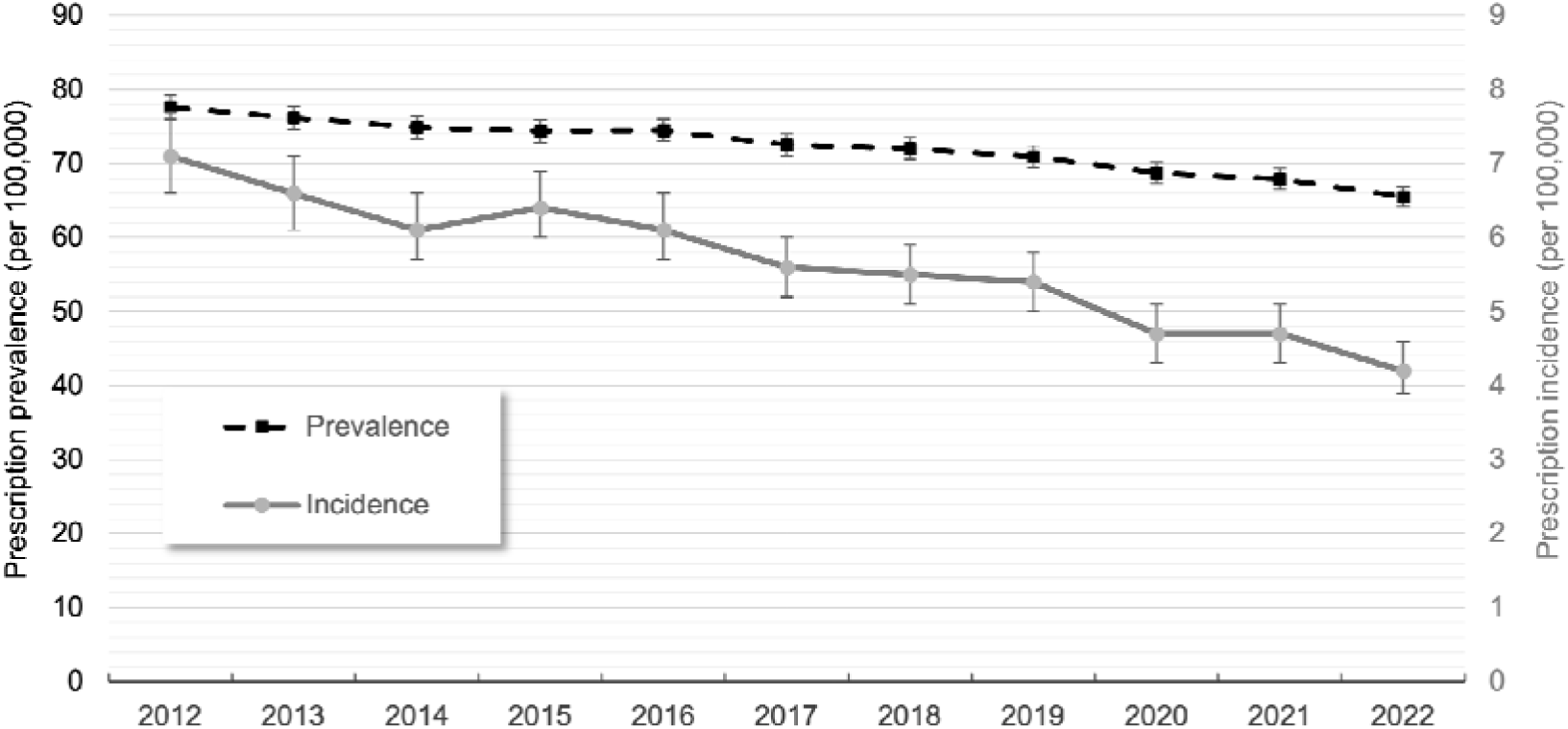
Overall age- and sex-standardized prescription prevalence and incidence (with 95% CIs) of clozapine between 2012 and 2022

Among females, in 2022, the prescription prevalence of clozapine increased with age, peaking at approximately 100 per 100,000 in the age groups from 50 to 64 years (**Figure 3A; Supplementary Table S1**). Between 2012 and 2022, the most pronounced relative declines in prevalence among women occurred in the age groups 30–34 (−51%) and 35–39 (−47%) years. Among males, in 2022, the prescription prevalence of clozapine also increased with age, peaking at approximately 160 per 100,000 in the age group 40–44 years (before declining in older age groups; **Figure 3B; Supplementary Table S1**). Between 2012 and 2022, the most pronounced relative declines in prevalence among men occurred in the age groups 26–29 (−48%) and 30–34 (−57%) years.

**Figure 3:**
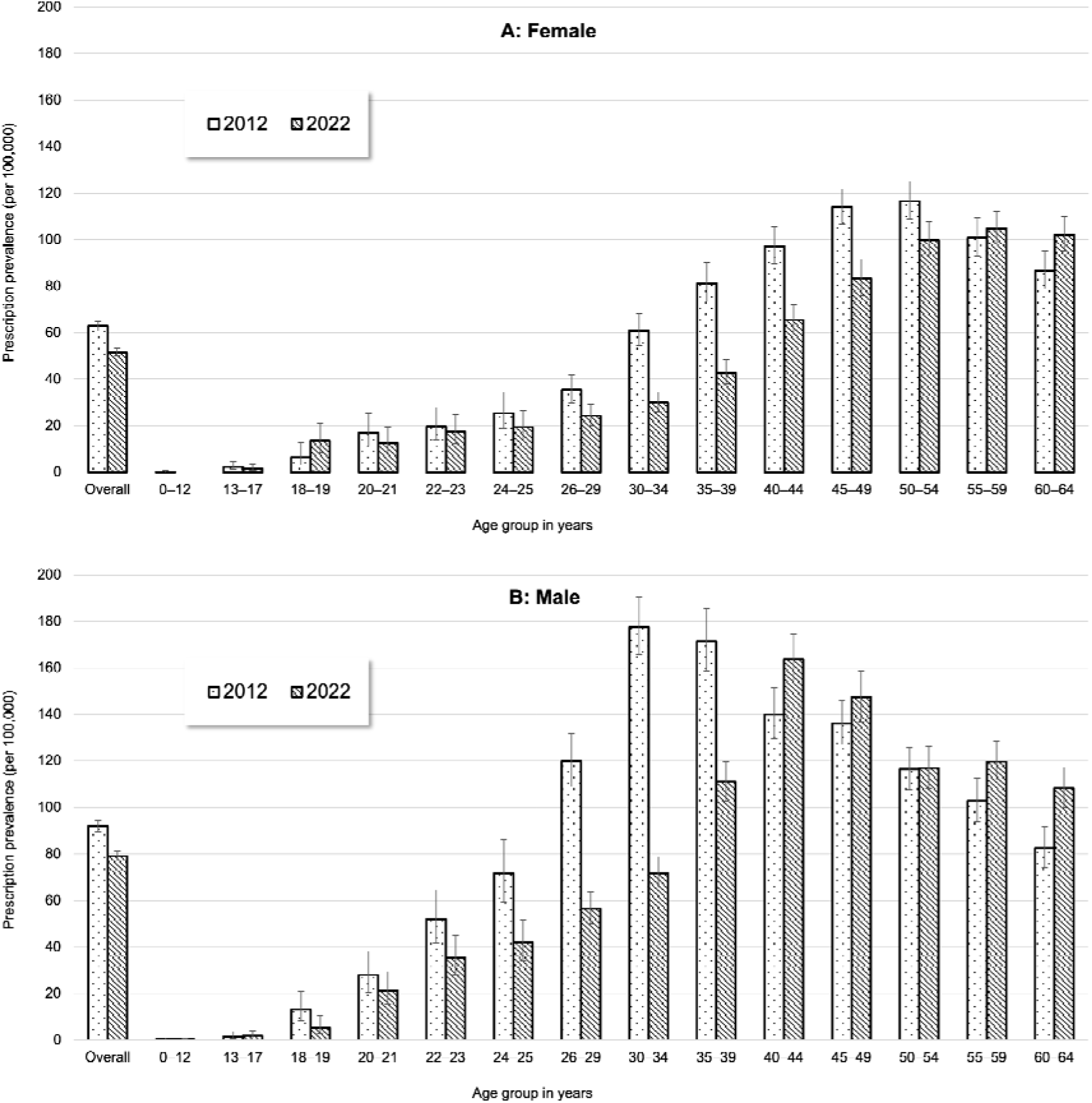
Prescription prevalence (with 95% CIs) of clozapine by age group among females (A) and males (B) in 2012 and 2022

### Temporal trends by urbanicity and socioeconomic deprivation, and district-level variation

In 2022, the age- and sex-standardized prescription prevalence of clozapine was highest in large urban cities at 73.1 per 100,000 (95% CI: 70.7; 75.7), compared to urban districts (63.0 per 100,000 [95% CI: 60.9; 65.2]), rural districts (62.4 per 100,000 [95% CI: 58.9; 66.1]), and sparsely populated rural districts (63.0 per 100,000 [95% CI: 59.2; 67.1]) (**Figure 4; Supplementary Table S3**). Between 2012 and 2022, the prescription prevalence declined in large urban cities (−23%) and urban districts (−16%) and remained largely stable in rural districts.

**Figure 4:**
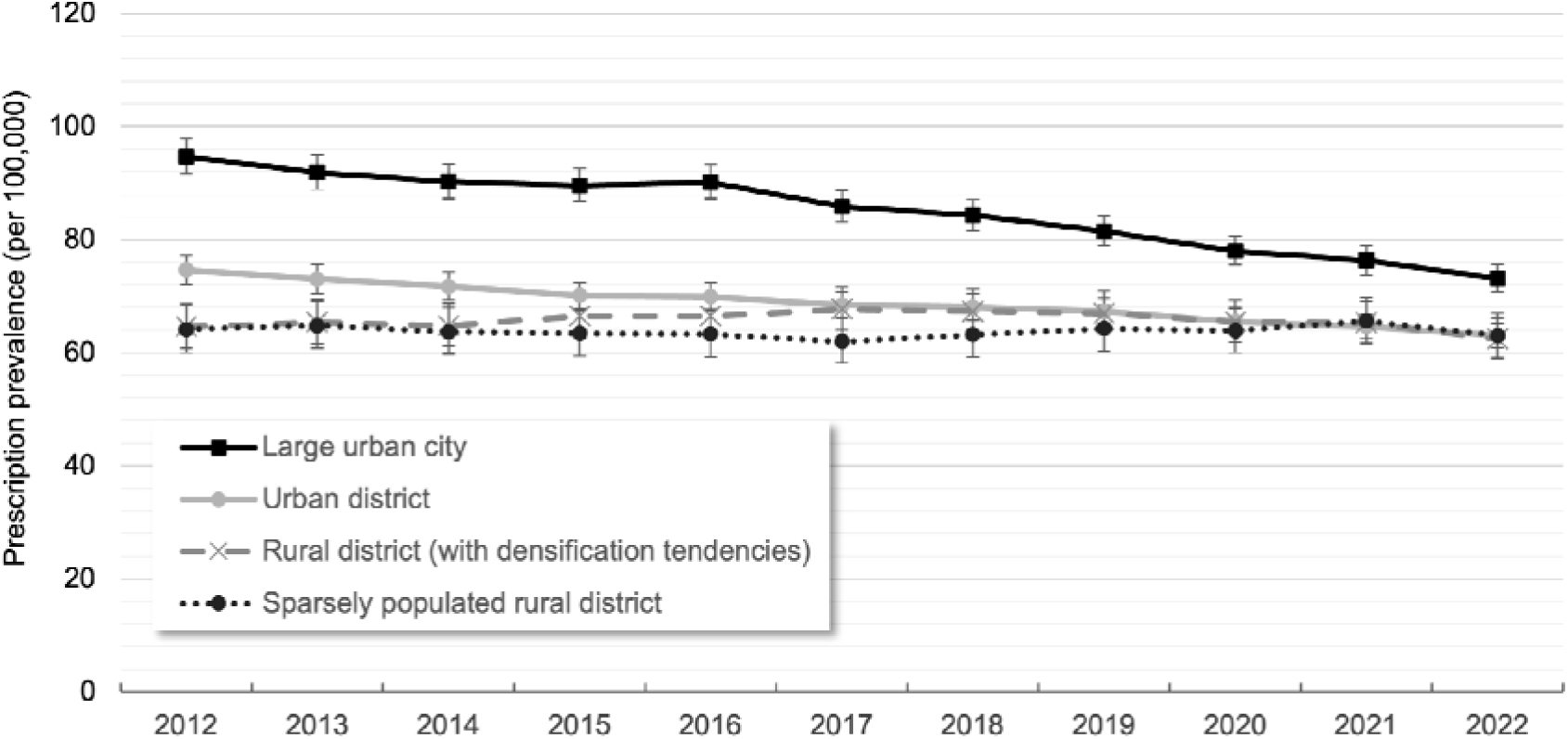
Age- and sex-standardized prescription prevalence (with 95% CIs) of clozapine by urbanicity between 2012 and 2022

Between 2012 and 2022, the age- and sex-standardized clozapine prescription prevalence declined across all strata of district-level socioeconomic deprivation, with the steepest decline observed in the least deprived areas. In the quintile representing the least deprived districts, the prevalence decreased by 22% (from 88.7 to 69.2 per 100,000). In the middle three quintiles (second to fourth), the decline was 14% (from 74.7 to 64.2 per 100,000), and in the most deprived quintile, it was 9% (from 72.6 to 66.3 per 100,000) (**Supplementary Figure S1 and Table S3**).

Of the 401 German districts, we included only those with a database population of ≥20,000 individuals in 2022 (n=202; **Table 1; Supplementary Table S4**). These districts account for approximately 73% of the total German population (BBSR, 2024). As indicated by the 5th and 95th percentile, the 5% of the districts with the lowest prescription prevalence of clozapine had values ≤29.7 per 100,000, while the top 5% had values ≥139.3 per 100,000, corresponding to a 4.7-fold difference. The full range spanned from 5.4 to 209 per 100,000, corresponding to a 38.7-fold difference in clozapine prescription prevalence between districts.

**Table 1:** Distribution of the standardized prescription prevalence of clozapine across 202 districts in 2022 (per 100,000 individuals)

| **Distribution statistics** | **Prevalence per 100,000**<br>**(n = 202 districts)** |
| --- | --- |
| Median | 57.9 |
| First quartile; third quartile | 42.3; 77.5 |
| 5th percentile; 95th percentile | 29.7; 139.3 |
| Minimum; maximum | 5.4; 209.0 |

Estimates of the prevalence proportion of clozapine prescriptions are age- and sex-standardized to the population of Germany as of December 31, 2022.
The sample included 202 (out of 401) districts with $\geq 20,000$ individuals in the database population. Based on the total population of these districts in official statistics, they represent approximately 73% of the overall German population in 2022.

## DISCUSSION

From 2012 to 2022, we found a decline in outpatient clozapine prescribing in Germany. This decline was mainly due to (a) a decrease in the number of individuals newly initiated on clozapine, (b) less prescribing among women aged 30–39 years and men aged 26–34 years, and (c) reduced prescribing in urban areas and in regions with higher socioeconomic status. In addition, we observed substantial regional variation in clozapine prescribing at the district level.

Our study is the first to update trends on the prescription prevalence of clozapine in a European country since the international analysis of 17 countries from 2005 to 2014 (Bachmann et al., 2017). In that study, clozapine prescription prevalence in Germany increased between 2005 and 2010, then plateaued at approximately 95 per 100,000 individuals until 2014. The authors concluded that clozapine remained underutilized in many countries and emphasized the need for targeted interventions to promote its use (Bachmann et al., 2017). Extending these findings, our study shows that clozapine use in Germany did not increase beyond 2014; instead, it declined steadily until 2022, with no indication of a new plateau.

Based on the same data source as the present study, a recent publication showed a continuous decrease in the prevalence of treated schizophrenia between 2012 and 2021 (−9%) (Riedel et al., 2025). Hence, the decrease in clozapine prescribing seen in the present study might at least partly be attributable to a decrease in schizophrenia prevalence in our population.

The estimated clozapine prescription prevalences in both studies presenting data for Germany (Bachmann et al. and ours) likely indicate clozapine underutilization, given that they are far below the expected prevalence estimates for TRS. With 12-month prevalence estimates of schizophrenia ranging from 330 to 460 per 100,000 in the general population (McGrath et al., 2008; Simeone et al., 2015) and 36.7% of all individuals with schizophrenia fulfilling TRS criteria (Diniz et al., 2023), the corresponding 12-month prevalence of TRS, i.e., the expected prescription prevalence of clozapine, can be estimated at approximately 121 to 169 per 100,000.

The lower clozapine prescription prevalence observed during the overlapping years 2012 to 2014 in our study (<80 per 100,000), compared to approximately 95 per 100,000 reported by Bachmann et al. (2017), may be attributed to two key differences: first, our study excluded individuals aged ≥65 years; second, there were differences in the socioeconomic characteristics—and therefore the underlying schizophrenia risk—of the study populations covered by the respective statutory health insurance providers (Hoffmann and Icks, 2012). Despite this discrepancy in overall prevalence, the sex-specific age patterns observed in our study closely mirror those reported by Bachmann et al. (2017) for Germany.

The more pronounced decline in clozapine prescribing incidence compared to the prevalence observed in our study indicates marked changes in prescribing patterns over the study period. First, substantially fewer individuals were newly started on clozapine, defined as clozapine initiation following a clozapine-free period of at least one year. Second, among those already receiving clozapine, the average treatment duration appears to have increased. This interpretation is supported by the established relationship between prevalence, incidence, and duration (Rothman and Greenland, 2023). If prescription incidence declines more sharply than prescription prevalence, this implies that treated individuals remain on treatment (here: clozapine) for longer, thereby slowing the decline in prevalence relative to incidence. The reasons for the decrease in clozapine initiation remain unclear, especially as the key position of clozapine in the treatment of TRS has continually been strengthened through clinical guidelines and quality assurance measures. Qualitative studies drawing on prescribers’ attitudes towards clozapine in TRS might help to elucidate the motives behind this lamentable trend.

To our knowledge, no study so far has examined the association between clozapine prescribing and urbanicity or socioeconomic deprivation in the general population. Among studies focusing specifically on patients with schizophrenia, clozapine use has been reported to be higher in urban areas compared to rural areas (Hou et al., 2019), while another study found no association with socioeconomic deprivation (Martin et al., 2014). However, these findings are not directly comparable to results based on general population data, as the risk of schizophrenia itself varies by both urbanicity (Vassos et al., 2012) and socioeconomic status (Agerbo et al., 2015; Werner et al., 2007). Notably, in contrast to the general risk of schizophrenia, higher urbanicity has been associated with a lower risk of TRS among patients with schizophrenia (Smart et al., 2021; Wimberley et al., 2016). Given that the design of these studies already accounts for regional differences in the risk of schizophrenia, these findings do not imply a higher clozapine prescribing prevalence in rural areas compared to urban areas in the general population. Regardless of the complex association of schizophrenia and TRS with urbanicity, our findings demonstrate that the observed overall decline in clozapine prescription prevalence is primarily seen in urban regions and those with higher socioeconomic status.

Several recent studies have examined regional variations in clozapine prescribing among general, i.e. unrestricted, populations in countries such as the United States (US) (Benito et al., 2023; Cavanah et al., 2025), the United Kingdom (UK) (Whiskey et al., 2021), Japan (Higuchi et al., 2022), and Norway (Schou et al., 2019). While these studies reported substantial intra-country regional differences in prescribing patterns, these analyses were based on relatively large administrative units, such as 50 states in the US, 14 regions in England, or 19 counties in Norway. Our study examined prescribing patterns at a much finer spatial resolution, thus revealing even more pronounced regional variations—up to a 38-fold difference. Compared with findings from other countries, lower regional differences have been reported: up to 13-fold in the US in 2019 (Benito et al., 2023), 2.4-fold in the UK (Whiskey et al., 2021), and 1.6-fold in Norway (Schou et al., 2019). Variation of this magnitude is unlikely to be attributable solely to differences in the prevalence or burden of schizophrenia. Rather, it indicates significant inconsistencies in prescribing practices which may reflect deviations from evidence-based care in the majority of studied regions.

### Strengths and limitations

A key strength of our study lies in the use of a large sample of routinely collected healthcare data covering a substantial proportion of the general population in Germany. This enabled comprehensive, population-based analyses, while minimizing recall and non-responder bias. Moreover, we did not restrict our study to patients with schizophrenia, even though clozapine underutilization in this patient population was the focus of our study. Nevertheless, by restricting our study population to people below 65 years of age, most patients who might have been prescribed clozapine for other indications (e.g., psychosis in Parkinson’s disease), were likely excluded.

However, our study also has several limitations. First, we could not capture clozapine prescriptions issued during inpatient stays. Consequently, our findings may underestimate the overall level of clozapine use, particularly around treatment initiation, which often occurs in hospital settings. Nonetheless, outpatient prescriptions reflect ongoing maintenance treatment and are thus highly relevant for assessing longitudinal trends and regional differences in routine clinical practice. Second, although this study was based on a large data base, we did not include the whole German population. Finally, no information was available on the duration of untreated psychosis. If this duration decreased over the study period, the number of persons with TRS—and therefore those eligible for clozapine treatment—would also have decreased, which could at least partly explain the observed decline in clozapine prescribing.

### Implications

Assessing population-based trends and regional variation in clozapine prescribing using healthcare data is essential for evaluating its use in routine clinical practice. Such real-world data has the potential to reveal the extent of underutilization and help determine whether prescribing practices align with clinical needs. Analyses stratified by demographic characteristics may elucidate the drivers of temporal trends, while identifying regions with low prescribing rates can uncover potential access barriers. These data can inform policymakers and guide the development of targeted interventions for adequate clozapine utilization.

Our findings underscore the need for effective measures to promote evidence-based prescribing of clozapine for people with TRS in Germany—a need that likely extends to many other countries as well. Barriers to more widespread use of clozapine appear to be predominantly prescriber-related (Agid et al., 2024; Bittner et al., 2023; Cohen, 2014; Cotes et al., 2022). This includes factors such as disproportionate concerns about safety and complexity of monitoring as well as systemic limitations and stigmatization of schizophrenia among mental health professionals (Bittner et al., 2023; Kane et al., 2019). Moreover, psychiatrists frequently anticipate low patient acceptance of clozapine (Cotes et al., 2022).

Increasing reluctance of prescribers to even offer clozapine could contribute to the decline in clozapine initiation in younger patients evident in our data. This is not only clinically unjustified but ethically problematic (John et al., 2018) as patients might often not even be adequately informed about clozapine as a vital treatment option (Kane, 2011; Rubio and Kane, 2021) despite clear evidence of high patient satisfaction, treatment preference, and adherence once it is initiated (van der Horst et al., 2025; Lieslehto et al., 2022; Parkes et al., 2022).

Emphasizing the patients’ perspective and needs through shared decision-making constitutes a critical yet underused tool that significantly improves the likelihood of clozapine recommendations and addresses misalignments between prescriber beliefs and patient preferences (Bittner et al., 2023; Falzer and Garman, 2012; Siskind et al., 2025). To this end, patient advocacy should also be crucial (Leung et al., 2023)—especially for reducing the considerable stigma associated with TRS (Kane et al., 2019)—but remains underinvestigated.

Perhaps most importantly, clozapine’s superior efficacy for reducing mortality in TRS should be sufficient to motivate its use (Bittner et al., 2023). Mortality reduction is an unequivocal treatment priority in other medical disciplines. Adopting this strategy for psychiatry should help to facilitate adequate clozapine utilization. To this end, introducing formalized and mandatory training regimes for psychiatrists regarding all aspects of clozapine use will be essential (Cohen and Farooq, 2020). This should be complemented by the systematic establishment of specialized TRS treatment teams (Bittner et al., 2023).

While our results suggest that treatment initiation might face the greatest barriers, successful clozapine initiation and its continued use also require adequate prevention and management of its side effects (Qubad and Bittner, 2023). Systemic barriers also need to be addressed. Strict blood monitoring requirements constitute an important barrier both from the patient and prescriber perspectives (Bittner et al., 2023). One important and recently realized measure has been a regulatory label change for clozapine by the European Medicines Agency (EMA), following longstanding demands from experts including the European Clozapine Task Force (Verdoux et al., 2025). The previous requirement of lifelong monthly blood monitoring was not evidence-based and increasingly viewed as outdated and burdensome (Meyer and Rubio, 2025). As of 2025, EMA recommends reducing the frequency of blood monitoring to every 12 weeks after the first treatment year without neutropenia, and to annual checks after two years, focusing solely on the absolute neutrophil count (ANC) rather than total leukocyte counts. This revision is expected to improve access to clozapine for individuals with TRS, reduce stigma and logistical barriers, and potentially save lives (Meyer and Rubio, 2025; Verdoux et al., 2025).

A recent global Delphi consensus aligns with these recommendations, advocating for lower ANC thresholds for clozapine cessation and discontinuation of routine absolute neutrophil count monitoring after two years (Siskind et al., 2025). The panel further emphasizes the need for quarterly comprehensive adverse drug reaction monitoring and simplified protocols to reduce patient burden. These evidence-based measures offer a more practical and patient-centered approach to clozapine safety, aiming to enhance outcomes for patients with TRS. Moreover, the reduced costs resulting from these changes should free resources urgently needed to improve other crucial aspects of care for patients with TRS (Diederich et al., 2025).

## CONCLUSIONS

Clozapine prescribing in Germany did not increase from 2012 to 2022, despite new clozapine-favoring guidelines, and showed substantial regional variation. Our results suggest a persisting underutilization of clozapine in most regions in Germany. Further research on barriers and facilitators on clozapine use in Germany is needed.

## STATEMENTS

### AUTHOR CONTRIBUTIONS

According to the CRediT taxonomy, OHFS contributed to Conceptualization, Methodology, Formal analysis, and Writing – original draft. OR, MQ, and MD contributed to Conceptualization, Methodology, and Writing – review & editing. BK contributed to Conceptualization, Methodology, Formal analysis, and Writing – review & editing. RAB and CJB contributed to Conceptualization, Methodology, and Writing – review & editing. All authors read and approved the final version of the manuscript.

### ETHICS STATEMENT

In Germany, the utilization of health insurance data for scientific research is regulated by the Code of Social Law. All involved health insurance providers as well as the Federal Office for Social Security and the Senator for Health, Women and Consumer Protection in Bremen as their responsible authorities approved the use of data for this study. Informed consent for studies based on claims data is required by law unless obtaining consent appears unacceptable and would bias results, which was the case in this study. According to the Ethics Committee of the University of Bremen studies based solely on pseudonymized personal data are exempt from institutional review board review.

### DATA AVAILABILITY STATEMENT

As we are not the owners of the data, we are not legally entitled to grant access to the data. In accordance with German data protection regulations, access to the data is granted only to employees of the Leibniz Institute for Prevention Research and Epidemiology – BIPS on the BIPS premises and in the context of approved research projects. Third parties may only access the data in cooperation with BIPS and after signing an agreement for guest researchers at BIPS.

## ACKNOWLEDGEMENTS

The authors would like to thank all statutory health insurance providers which provided data for this study, namely AOK Bremen/Bremerhaven, DAK-Gesundheit, Techniker Krankenkasse (TK), and hkk Krankenkasse. They would also like to thank Alina Ludewig and Fabian Gesing for the statistical programming of the data and Dr. Heike Gerds for proofreading the final manuscript (all with Leibniz Institute – BIPS).

## REFERENCES

Agerbo, E., Sullivan, P.F., Vilhjálmsson, B.J., Pedersen, C.B., Mors, O., Børglum, A.D., Hougaard, D.M., Hollegaard, M.V., Meier, S., Mattheisen, M., Ripke, S., Wray, N.R., Mortensen, P.B., 2015. Polygenic Risk Score, Parental Socioeconomic Status, Family History of Psychiatric Disorders, and the Risk for Schizophrenia: A Danish PopulationBased Study and Meta-analysis. JAMA Psychiatry 72, 635–641. 10.1001/jamapsychiatry.2015.0346

Agid, O., Crespo-Facorro, B., Bartolomeis, A. de, Fagiolini, A., Howes, O.D., Seppälä, N., Correll, C.U., 2024. Overcoming the barriers to identifying and managing treatment-resistant schizophrenia and to improving access to clozapine: A narrative review and recommendation for clinical practice. Eur. Neuropsychopharmacol. 84, 35–47. 10.1016/j.euroneuro.2024.04.012

Bachmann, C.J., Aagaard, L., Bernardo, M., Brandt, L., Cartabia, M., Clavenna, A., Fusté, A.C., Furu, K., Garuoliené, K., Hoffmann, F., Hollingworth, S., Huybrechts, K.F., Kalverdijk, L.J., Kawakami, K., Kieler, H., Kinoshita, T., López, S.C., Machado□Alba, J.E., Machado□Duque, M.E., Mahesri, M., Nishtala, P.S., Piovani, D., Reutfors, J., Saastamoinen, L.K., Sato, I., Schuiling□Veninga, C.C.M., Shyu, Y. C., Siskind, D., Skurtveit, S., Verdoux, H., Wang, L. □J., Yahni, C.Z., Zoëga, H., Taylor, D., 2017. International trends in clozapine use: a study in 17 countries. Acta Psychiat Scand 136, 37–51. 10.1111/acps.12742

BBSR, 2024. Indikatoren und Karten zur Raum- und Stadtentwicklung. INKAR. Ausgabe 2024. Hrsg.: Bundesinstitut für Bau-, Stadt- und Raumforschung (BBSR) im Bundesamt für Bauwesen und Raumordnung (BBR), Bonn [WWW Document]. URL https://www.inkar.de (accessed 6.24.25).

Benito, R.A., Gatusky, M.H., Panoussi, M.W., McCall, K.L., Suparmanian, A.S., Piper, B.J., 2023. Thirteen-fold variation between states in clozapine prescriptions to United States Medicaid patients. Schizophr. Res. 255, 79–81. 10.1016/j.schres.2023.03.010

Bittner, R.A., Reif, A., Qubad, M., 2023. The ever-growing case for clozapine in the treatment of schizophrenia: an obligation for psychiatrists and psychiatry. Curr. Opin. Psychiatry 36, 327–336. 10.1097/yco.0000000000000871

Brodeur, S., Courteau, J., Vanasse, A., Courteau, M., Stip, E., Fleury, M.-J., Lesage, A., Demers, M.-F., Corbeil, O., Béchard, L., Roy, M.-A., 2022. Association between previous and future antipsychotic adherence in patients initiating clozapine: real-world observational study. Br. J. Psychiatry 220, 347–354. 10.1192/bjp.2022.1

Brunette, M.F., Drake, R.E., Xie, H., McHugo, G.J., Green, A.I., 2006. Clozapine use and relapses of substance use disorder among patients with co-occurring schizophrenia and substance use disorders. Schizophr. Bull. 32, 637–643. 10.1093/schbul/sbl003

Cavanah, L.R., Tian, M.Y., Goldhirsh, J.L., Huey, L.Y., Piper, B.J., 2025. Declines and pronounced state-level variation in clozapine use among Medicare patients. PLOS One 20, e0328495. 10.1371/journal.pone.0328495

Chakos, M., Lieberman, J., Hoffman, E., Bradford, D., Sheitman, B., 2001. Effectiveness of second-generation antipsychotics in patients with treatment-resistant schizophrenia: a review and meta-analysis of randomized trials. Am. J. Psychiatry 158, 518–526. 10.1176/appi.ajp.158.4.518

Cohen, D., 2014. Prescribers fear as a major side □effect of clozapine. Acta Psychiatr. Scand. 130, 154–155. 10.1111/acps.12294

Cohen, D., Farooq, S., 2020. Mandatory certification for clozapine prescribing. Eur. Psychiatry 64, e12. 10.1192/j.eurpsy.2020.110

Correll, C.U., Martin, A., Patel, C., Benson, C., Goulding, R., Kern-Sliwa, J., Joshi, K., Schiller, E., Kim, E., 2022a. Systematic literature review of schizophrenia clinical practice guidelines on acute and maintenance management with antipsychotics. Schizophrenia 8, 5. 10.1038/s41537-021-00192-x

Correll, C.U., Solmi, M., Croatto, G., Schneider, L.K., Rohani□Montez, S.C., Fairley, L., Smith, N., Bitter, I., Gorwood, P., Taipale, H., Tiihonen, J., 2022b. Mortality in people with schizophrenia: a systematic review and meta□analysis of relative risk and aggravating or attenuating factors. World Psychiatry 21, 248–271. 10.1002/wps.20994

Cotes, R.O., Janjua, A.U., Broussard, B., Lazris, D., Khan, A., Jiao, Y., Kopelovich, S.L., Goldsmith, D.R., 2022. A comparison of attitudes, comfort, and knowledge of clozapine among two diverse samples of US psychiatrists. Community Ment. Heal. J. 58, 517–525. 10.1007/s10597-021-00847-0

Deutsche Gesellschaft für Psychiatrie und Psychotherapie, Psychosomatik und Nervenheilkunde, 2025. S3-Leitlinie Schizophrenie – Living Guideline (AWMF Registry No. 038-009; Version 4.0). Association of the Scientific Medical Societies in Germany (AWMF). [WWW Document]. URL https://register.awmf.org/de/leitlinien/detail/038-009 (accessed 11.19.25).

Diederich, F., Oloyede, E., Taylor, D., Bachmann, C.J., 2025. Economic evaluation of different haematological monitoring schemes for patients with treatment-resistant schizophrenia using clozapine. Br. J. Psychiatry Published online, 1–8. 10.1192/bjp.2025.10424

Diniz, E., Fonseca, L., Rocha, D., Trevizol, A., Cerqueira, R., Ortiz, B., Brunoni, A.R., Bressan, R., Correll, C.U., Gadelha, A., 2023. Treatment resistance in schizophrenia: a meta-analysis of prevalence and correlates. Braz. J. Psychiatry 45, 448–458. 10.47626/1516-4446-2023-3126

Faay, M.D.M., Czobor, P., Sommer, I.E.C., 2018. Efficacy of typical and atypical antipsychotic medication on hostility in patients with psychosis-spectrum disorders: a review and meta-analysis. Neuropsychopharmacol 43, 2340–2349. 10.1038/s41386-018-0161-2

Falzer, P.R., Garman, D.M., 2012. Optimizing clozapine through clinical decision making. Acta Psychiatr. Scand. 126, 47–58. 10.1111/j.1600-0447.2012.01863.x

Frogley, C., Taylor, D., Dickens, G., Picchioni, M., 2012. A systematic review of the evidence of clozapine’s anti-aggressive effects. Int. J. Neuropsychopharmacol. 15, 1351–1371. 10.1017/s146114571100201x

Haug, U., Schink, T., 2021. German Pharmacoepidemiological Research Database (GePaRD), in: Sturkenboom, M., Schink, T. (Eds.), Databases for Pharmacoepidemiological Research, Springer Series on Epidemiology and Public Health. Springer, Cham, Switzlerland, pp. 119–124. 10.1007/978-3-030-51455-6_8

Hennen, J., Baldessarini, R.J., 2005. Suicidal risk during treatment with clozapine: a metaanalysis. Schizophr. Res. 73, 139–145. 10.1016/j.schres.2004.05.015

Higuchi, S., Sako, A., Kondo, T., Kusanishi, S., Enomoto, T., Hayakawa, T., Yanai, H., Yoshimura, K., 2022. Clozapine use in Japan based on national database of health insurance claims and specific health checkups open data: disparities by region, age, and sex. Psychiatria et Neurologia Japonica 124, 3–15.

Hoffmann, F., Icks, A., 2012. [Structural differences between health insurance funds and their impact on health services research: results from the Bertelsmann Health-Care Monitor]. Gesundheitswesen 74, 291–297. 10.1055/s-0031-1275711

Horst, M.Z. van der, Boer, N. de, Müderrisoğlu, A., Privat, A.T., Bouhuis, A., Hasan, A., Jongkind, A., Gonzalez-Pinto, A., Santacana, A.M., D’Agostino, A., Ertuğrul, A., Yağcıoğlu, A.E.A., Crespo-Facorro, B., Baune, B.T., Sanchez-Barbero, B., Spuch, C., Morgenroth, C.L., Pinedo, C.F. de, Casetta, C., Bousman, C., Hohoff, C., Pantelis, C., Ovejas-Catalán, C., Garcia-Rizo, C., Cohen, D., Ristic, D.I., Beld, E., Repo-Tiihonen, E., Wagner, E., Jeger-Land, E., Vilella, E., Fernandez-Egea, E., Bekema, E., Sepúlveda, E., Wiedenmann, F., Martini, F., Serio, F., Vairano, F., Mercuriali, G., Boido, G., Yoca, G., Beek, H. van, Gijsman, H., Tuppurainen, H., Everall, I., Novakovic, I., Zorrilla, I., Erdoğan, İ.M., Sapienza, J., Bogers, J., Tiihonen, J., Vázquez-Bourgon, J., Os, J. van, Schneider-Thoma, J., Luykx, J., Grootens, K., Mar-Barrutia, L., Martorell, L., Bak, M., Spangaro, M., Zierhut, M., Vos, M. de, Koning, M. de, Garriga, M., Lähteenvuo, M., Bosia, M., Horst, M. van der, Babaoğlu, M.O., Veereschild, M., Manchia, M., Qubad, M., Edlinger, M., Fuentes-Pérez, P., Paribello, P., Lopez-Pena, P., Kahn, R., Bittner, R.A., Cavallaro, R., Veerman, S., Gutwinski, S., Schreiter, S., Ripke, S., Baltanás, T.R., Oviedo-Salcedo, T., Hallikainen, T., Görlitz, T., Alink, W., Ayhan, Y., Okhuijsen-Pfeifer, C., Okhuijsen-Pfeifer, C., Luykx, J.J., 2025. Determinants of patient satisfaction in clozapine users: results from the Clozapine International Consortium (CLOZIN). Schizophrenia 11, 28. 10.1038/s41537-025-00570-9

Hou, C.-L., Wang, S.-B., Wang, F., Xu, M.-Z., Chen, M.-Y., Cai, M.-Y., Xiao, Y.-N., Jia, F.-J., 2019. Psychotropic medication treatment patterns in community-dwelling schizophrenia in China: comparisons between rural and urban areas. BMC Psychiatry 19, 242. 10.1186/s12888-019-2217-1

Howes, O.D., McCutcheon, R., Agid, O., Bartolomeis, A. de, Beveren, N.J.M. van, Birnbaum, M.L., Bloomfield, M.A.P., Bressan, R.A., Buchanan, R.W., Carpenter, W.T., Castle, D.J., Citrome, L., Daskalakis, Z.J., Davidson, M., Drake, R.J., Dursun, S., Ebdrup, B.H., Elkis, H., Falkai, P., Fleischacker, W.W., Gadelha, A., Gaughran, F., Glenthøj, B.Y., Graff-Guerrero, A., Hallak, J.E.C., Honer, W.G., Kennedy, J., Kinon, B.J., Lawrie, S.M., Lee, J., Leweke, F.M., MacCabe, J.H., McNabb, C.B., Meltzer, H., Möller, H.-J., Nakajima, S., Pantelis, C., Marques, T.R., Remington, G., Rossell, S.L., Russell, B.R., Siu, C.O., Suzuki, T., Sommer, I.E., Taylor, D., Thomas, N., Üçok, A., Umbricht, D., Walters, J.T.R., Kane, J., Correll, C.U., 2017. Treatment-Resistant Schizophrenia: Treatment Response and Resistance in Psychosis (TRRIP) Working Group Consensus Guidelines on Diagnosis and Terminology. Am. J. Psychiatry 174, 216–229. 10.1176/appi.ajp.2016.16050503

Huhn, M., Nikolakopoulou, A., Schneider-Thoma, J., Krause, M., Samara, M., Peter, N., Arndt, T., Bäckers, L., Rothe, P., Cipriani, A., Davis, J., Salanti, G., Leucht, S., 2019. Comparative efficacy and tolerability of 32 oral antipsychotics for the acute treatment of adults with multi-episode schizophrenia: a systematic review and network meta-analysis. Lancet 394, 939–951. 10.1016/s0140-6736(19)31135-3

John, A.P., Ko, E.K.F., Dominic, A., 2018. Delayed initiation of clozapine continues to be a substantial clinical concern. Can. J. Psychiatry 63, 526–531. 10.1177/0706743718772522

Jones, R., Upthegrove, R., Price, M.J., Pritchard, M., Chandan, J.S., Legge, S., MacCabe, J.H., 2022. Duration of prior psychotic illness and clozapine response: a retrospective observational study using electronic health records. Ther. Adv. Psychopharmacol. 12, 20451253221103353. 10.1177/20451253221103353

Kane, J., Honigfeld, G., Singer, J., Meltzer, H., 1988. Clozapine for the treatment-resistant schizophrenic: a double-blind comparison with chlorpromazine. Arch. Gen. Psychiatry 45, 789–796. 10.1001/archpsyc.1988.01800330013001

Kane, J.M., 2011. A user’ guide to clozapine. Acta Psychiatr. Scand. 123, 407–408. 10.1111/j.1600-0447.2011.01711.x

Kane, J.M., Agid, O., Baldwin, M.L., Howes, O., Lindenmayer, J.-P., Marder, S., Olfson, M., Potkin, S.G., Correll, C.U., 2019. Clinical Guidance on the Identification and Management of Treatment-Resistant Schizophrenia. J. Clin. Psychiatry 80. 10.4088/jcp.18com12123

Lähteenvuo, M., Batalla, A., Luykx, J.J., Mittendorfer□Rutz, E., Tanskanen, A., Tiihonen, J., Taipale, H., 2021. Morbidity and mortality in schizophrenia with comorbid substance use disorders. Acta Psychiatr. Scand. 144, 42–49. 10.1111/acps.13291

Lähteenvuo, M., Luykx, J.J., Taipale, H., Mittendorfer-Rutz, E., Tanskanen, A., Batalla, A., Tiihonen, J., 2022. Associations between antipsychotic use, substance use and relapse risk in patients with schizophrenia: real-world evidence from two national cohorts. Br. J. Psychiatry 221, 758–765. 10.1192/bjp.2022.117

Leung, J.G., Ehret, M., Love, R.C., Cotes, R.O., 2023. Improving clozapine utilization will require continued advocacy, drug sponsor interest, and FDA support to address REMS issues. Expert Rev. Clin. Pharmacol. 16, 177–179. 10.1080/17512433.2023.2183192

Lieslehto, J., Tiihonen, J., Lähteenvuo, M., Tanskanen, A., Taipale, H., 2022. Primary nonadherence to antipsychotic treatment among persons with schizophrenia. Schizophr. Bull. 48, 655–663. 10.1093/schbul/sbac014

Luykx, J.J., Colgan, M., Vieta, E., Hamina, A., Schulte, P.F.J., Correll, C.U., Mittendorfer-Rutz, E., Siskind, D., Lieslehto, J., Tanskanen, A., Tiihonen, J., Taipale, H., 2025. Transdiagnostic effectiveness and safety of clozapine in individuals with psychotic, affective, and personality disorders: nationwide and meta-analytic comparisons with other antipsychotics. Lancet Psychiatry 12, 921–931. 10.1016/s22150366(25)00297-4

Martin, D.J., Park, J., Langan, J., Connolly, M., Smith, D.J., Taylor, M., 2014. Socioeconomic status and prescribing for schizophrenia: analysis of 3200 cases from the Glasgow Psychosis Clinical Information System (PsyCIS). Psychiatr. Bull. 38, 54–57. 10.1192/pb.bp.112.042143

Masdrakis, V.G., Baldwin, D.S., 2023. Prevention of suicide by clozapine in mental disorders: systematic review. Eur. Neuropsychopharmacol. 69, 4–23. 10.1016/j.euroneuro.2022.12.011

Masuda, T., Misawa, F., Takase, M., Kane, J.M., Correll, C.U., 2019. Association With Hospitalization and All-Cause Discontinuation Among Patients With Schizophrenia on Clozapine vs Other Oral Second-Generation Antipsychotics. JAMA Psychiatry 76, 1052–1062. 10.1001/jamapsychiatry.2019.1702

McCutcheon, R.A., Pillinger, T., Varvari, I., Halstead, S., Ayinde, O.O., Crossley, N.A., Correll, C.U., Hahn, M., Howes, O.D., Kane, J.M., Kabir, T., Konradsson-Geuken, Å., Lennox, B., Hui, C.L.M., Rossell, S.L., Solmi, M., Sommer, I.E., Taipale, H., Uchida, H., Venkatasubramanian, G., Warren, N., Group, T.I.A., Agid, O., Arango, C., Bartolomeis, A. de, Bittner, R.A., Bressan, R.A., Buchanan, R.W., Cecere, G., Chen, E.Y.H., Citrome, L., Dollfus, S., Dursun, S.M., Ebdrup, B.H., Elkis, H., Emsley, R., Fernandez-Egea, E., Fleischhacker, W., Freudenreich, O., Gadelha, A., Gangadin, S.S., Gaughran, F., Hallak, J.E.C., Hasan, A., Homan, P., Jauhar, S., Kaiser, S., Kwon, J.S., Lee, J., Leucht, S., Libiger, J., Leweke, M.F., MacCabe, J.H., Marder, S.R., Marques, T.R., Melle, I., Mucci, A., Nakajima, S., O’Donoghue, B., Ojagbemi, A., Perry, B.I., Roos, J.L., Rubio, J.M., Rybakowski, J.K., Sachs, G., Siu, C.O., Suzuki, T., Taylor, D.M., Takeuchi, H., Thirthalli, J., Thomas, N., Üçok, A., Varambally, S., Vasiliu, O., Wagner, E., Siskind, D., 2025. INTEGRATE: international guidelines for the algorithmic treatment of schizophrenia. Lancet Psychiatry. 10.1016/s2215-0366(25)00031-8

McGrath, J., Saha, S., Chant, D., Welham, J., 2008. Schizophrenia: A Concise Overview of Incidence, Prevalence, and Mortality. Epidemiologic Rev. 30, 67–76. 10.1093/epirev/mxn001

Meltzer, H.Y., 2012. Clozapine: balancing safety with superior antipsychotic efficacy. Clin. Schizophr. Relat. Psychoses 6, 134–144. 10.3371/csrp.6.3.5

Meltzer, H.Y., Alphs, L., Green, A.I., Altamura, A.C., Anand, R., Bertoldi, A., Bourgeois, M., Chouinard, G., Islam, M.Z., Kane, J., Krishnan, R., Lindenmayer, J.P., Potkin, S., Group, I.S.P.T.S., 2003. Clozapine treatment for suicidality in schizophrenia: International Suicide Prevention Trial (InterSePT). Arch. Gen. Psychiatry 60, 82–91. 10.1001/archpsyc.60.1.82

Meyer, J.M., Rubio, J.M., 2025. Clozapine Monitoring in the Post-REMS World. J. Clin. Psychiatry 86. 10.4088/jcp.25ac15898

Michalski, N., Reis, M., Tetzlaff, F., Nowossadeck, E., Hoebel, J., 2022. German Index of Socioeconomic Deprivation (GISD), Berlin: Zenodo. [WWW Document]. URL 10.5281/zenodo.7973846 (accessed 12.1.23).

Mizuno, Y., McCutcheon, R.A., Brugger, S.P., Howes, O.D., 2020. Heterogeneity and efficacy of antipsychotic treatment for schizophrenia with or without treatment resistance: a meta-analysis. Neuropsychopharmacology 45, 622–631. 10.1038/s41386019-0577-3

Parkes, S., Mantell, B., Oloyede, E., Blackman, G., 2022. Patients’ experiences of clozapine for treatment-resistant schizophrenia: a systematic review. Schizophr. Bull. Open 3, sgac042. 10.1093/schizbullopen/sgac042

Qubad, M., Bittner, R.A., 2023. Second to none: rationale, timing, and clinical management of clozapine use in schizophrenia. Ther. Adv. Psychopharmacol. 13, 20451253231158152. 10.1177/20451253231158152

Riedel, O., Bachmann, C.J., Bittner, R.A., Dörks, M., Kollhorst, B., Qubad, M., Scholle, O.H.F., 2025. Prevalence and incidence of treated schizophrenia: temporal and regional trends in Germany. Schizophrenia 11, 131. 10.1038/s41537-025-00689-9

Rothman, K.J., Greenland, S., 2023. Basic Concepts, in: Ahrens, W., Pigeot, I. (Eds.), Handbook of Epidemiology. Springer, New York, NY. 10.1007/978-14614-6625-3_44-1

Rubio, J.M., Kane, J.M., 2021. How to make an effective offer of clozapine. J. Clin. Psychiatry 83. 10.4088/jcp.21ac14000

Schou, M.B., Drange, O.K., Sæther, S.G., 2019. Fylkesvise forskjeller i forskrivning av klozapin. Tidsskr. Den Nor. legeforening 139. 10.4045/tidsskr.19.0151

Shah, P., Iwata, Y., Brown, E.E., Kim, J., Sanches, M., Takeuchi, H., Nakajima, S., Hahn, M., Remington, G., Gerretsen, P., Graff-Guerrero, A., 2020. Clozapine response trajectories and predictors of non-response in treatment-resistant schizophrenia: a chart review study. Eur. Arch. Psychiatry Clin. Neurosci. 270, 11–22. 10.1007/s00406-019-01053-6

Simeone, J.C., Ward, A.J., Rotella, P., Collins, J., Windisch, R., 2015. An evaluation of variation in published estimates of schizophrenia prevalence from 1990─2013: a systematic literature review. BMC Psychiatry 15, 193. 10.1186/s12888015-0578-7

Siris, S.G., 2001. Suicide and schizophrenia. J. Psychopharmacol. 15, 127–135. 10.1177/026988110101500209

Siskind, D., Northwood, K., Pillinger, T., Chan, S., Correll, C., Cotes, R.O., Every-Palmer, S., Hahn, Maggie, Howes, O.D., Kane, J.M., Kelly, D., Korman, N., Lappin, J., Mena, C., Myles, Nick, McCutcheon, R.A., Panel, C.D.E., Siskind, D., McCutcheon, R.A., Northwood, K., Pillinger, T., Chan, S., Correll, C.U., Cotes, R., Every-Palmer, S., Hahn, Margaret, Howes, O., Kane, J.M., Kelly, D.L., Korman, N., Lappin, J., Mena, C., Myles, Nicholas, Agid, O., Arango, C., Asmal, L., Ayinde, O.O., Baptiste, T., Béchard, L., Bitter, I., Bittner, R.A., Breznoscakova, D., Chen, E., Chkonia, E., Citrome, L., Clark, S.R., Corbeil, O., Cordoba, R., Crossley, N., Bartolomeis, A. de, Ebdrup, B.H., Fernandez-Egea, E., Elkis, H., Eltorki, Y., Filipčić, I.Š., Filippis, R. de, Fleischhacker, W., Freudenreich, O., Gadelha, A., Gama, C.S., Gaughran, F., Gee, S., Guinart, D., Hallak, J.E.C., Harris, A., Hasan, A., Honer, W., Jayaram, M., Kanazawa, T., Kawashima, H., Keating, D., Kim, S.H., Kwon, J.S., Lane, H.-Y., Lee, J., Lennox, B., Leon, J. de, Leung, J.G., Luykx, J.J., MacCabe, J., Murray, G., Nielsen, J., O’Donoghue, B., Oloyede, E., Qubad, M., Remington, G., Roy, M.-A., Rubio, J., Sagud, M., Schulte, P.F.J., Selten, J.-P., Si, T., Smith, L., Sommer, I., Sundram, S., Taipale, H., Takeuchi, H., Tandon, R., Tanzer, T., Taylor, D., Thirthalli, J., Tiihonen, J., Uchida, H., Venkatasubramanian, G., Verdoux, H., Wagner, E., Xin, Y., Yağcıoğlu, A.E.A., Yung, A., 2025. Absolute neutrophil count and adverse drug reaction monitoring during clozapine treatment: consensus guidelines from a global Delphi panel. Lancet Psychiatry. 10.1016/s2215-0366(25)00098-7

Siskind, D., Orr, S., Sinha, S., Yu, O., Brijball, B., Warren, N., MacCabe, J.H., Smart, S.E., Kisely, S., 2022. Rates of treatment-resistant schizophrenia from first-episode cohorts: systematic review and meta-analysis. Br. J. Psychiatry 220, 115–120. 10.1192/bjp.2021.61

Smart, S.E., Kępińska, A.P., Murray, R.M., MacCabe, J.H., 2021. Predictors of treatment resistant schizophrenia: a systematic review of prospective observational studies. Psychol. Med. 51, 44–53. 10.1017/s0033291719002083

Solmi, M., Tiihonen, J., Lähteenvuo, M., Tanskanen, A., Correll, C.U., Taipale, H., 2021. Antipsychotics Use Is Associated With Greater Adherence to Cardiometabolic Medications in Patients With Schizophrenia: Results From a Nationwide, Within-subject Design Study. Schizophr. Bull. 48, 166–175. 10.1093/schbul/sbab087

Taipale, H., Tanskanen, A., Howes, O., Correll, C.U., Kane, J.M., Tiihonen, J., 2025. Comparative effectiveness of antipsychotic treatment strategies for relapse prevention in first-episode schizophrenia in Finland: a population-based cohort study. Lancet Psychiatry 12, 122–130. 10.1016/s2215-0366(24)00366-3

Taipale, H., Tanskanen, A., Mehtälä, J., Vattulainen, P., Correll, C.U., Tiihonen, J., 2020. 20□year follow□up study of physical morbidity and mortality in relationship to antipsychotic treatment in a nationwide cohort of 62,250 patients with schizophrenia (FIN20). World Psychiatry 19, 61–68. 10.1002/wps.20699

Vassos, E., Pedersen, C.B., Murray, R.M., Collier, D.A., Lewis, C.M., 2012. Meta-Analysis of the Association of Urbanicity With Schizophrenia. Schizophr. Bull. 38, 1118–1123. 10.1093/schbul/sbs096

Verdoux, H., Bittner, R.A., Hasan, A., Qubad, M., Wagner, E., Lepetit, A., Arrojo-Romero, M., Bachmann, C., Beex-Oosterhuis, M., Bogers, J., Celofiga, A., Cohen, D., Berardis, D. de, Hert, M. de, Cuevas, C. de L., Ebdrup, B.H., Fountoulakis, K.N., Guinart, D., Keating, D., Kopeček, M., Lally, J., Lazáry, J., Luykx, J.J., Amigo, O.M., Molden, E., Nielsen, J., O’Donoghue, B., Oswald, P., Radulescu, F.S., Rohde, C., Sagud, M., Sanz, E.J., Filipčić, I.Š., Sommer, I.E., Taipale, H., Tiihonen, J., Tuppurainen, H., Veerman, S., Wilkowska, A., Spina, E., Schulte, P., 2025. The time has come for revising the rules of clozapine blood monitoring in Europe. A joint expert statement from the European Clozapine Task Force. Eur. Psychiatry 68, e17. 10.1192/j.eurpsy.2024.1816

Wagner, E., Kane, J.M., Correll, C.U., Howes, O., Siskind, D., Honer, W.G., Lee, J., Falkai, P., Schneider-Axmann, T., Hasan, A., Group, T.W., 2020. Clozapine combination and augmentation strategies in patients with schizophrenia—recommendations from an international expert survey among the Treatment Response and Resistance in Psychosis (TRRIP) Working Group. Schizophr. Bull. 46, 1459–1470. 10.1093/schbul/sbaa060

Wagner, E., Siafis, S., Fernando, P., Falkai, P., Honer, W.G., Röh, A., Siskind, D., Leucht, S., Hasan, A., 2021. Efficacy and safety of clozapine in psychotic disorders—a systematic quantitative meta-review. Transl. Psychiatry 11, 487. 10.1038/s41398-02101613-2

Werner, S., Malaspina, D., Rabinowitz, J., 2007. Socioeconomic Status at Birth Is Associated With Risk of Schizophrenia: Population-Based Multilevel Study. Schizophr. Bull. 33, 1373–1378. 10.1093/schbul/sbm032

Whiskey, E., Barnard, A., Oloyede, E., Dzahini, O., Taylor, D.M., Shergill, S.S., 2021. An evaluation of the variation and underuse of clozapine in the United Kingdom. Acta Psychiatr. Scand. 143, 339–347. 10.1111/acps.13280

Wimberley, T., Støvring, H., Sørensen, H.J., Horsdal, H.T., MacCabe, J.H., Gasse, C., 2016. Predictors of treatment resistance in patients with schizophrenia: a population-based cohort study. Lancet Psychiatry 3, 358–366. 10.1016/s22150366(15)00575-1

Yada, Y., Yoshimura, B., Kishi, Y., 2015. Correlation between delay in initiating clozapine and symptomatic improvement. Schizophr. Res. 168, 585–586. 10.1016/j.schres.2015.07.045

