## Supplementary Material for "Clozapine prescribing in Germany: temporal trends and regional variations, 2012–2022"

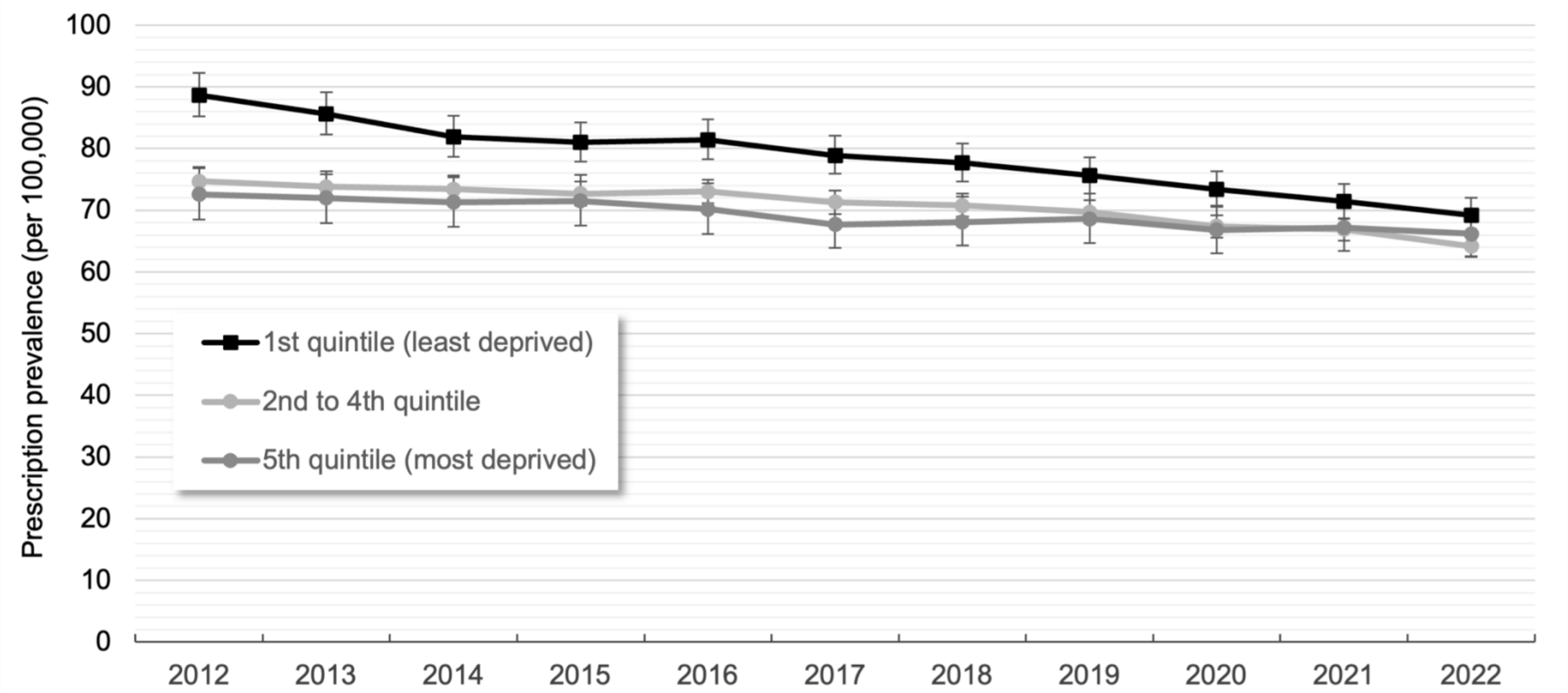

**Figure S1:** Age- and sex-standardized prescription prevalence (with 95% CIs) of clozapine by district-level socioeconomic deprivation between 2012 and 2022

**Table S1:** Standardized prescription prevalence of clozapine by age and sex for each calendar year from 2012 to 2022 (per 100,000 persons; 95% confidence intervals in brackets)

| | **2012** | **2013** | **2014** | **2015** | **2016** |
|---|---|---|---|---|---|
| **Total number of (database) population, 0–64 years, n** | 11,653,312 | 12,133,011 | 12,374,789 | 12,661,181 | 12,735,204 |
| Total number of (database) population, all age years, n | 14,677,493 | 15,241,994 | 15,570,274 | 15,948,015 | 16,079,136 |
| Overall, all age years | 81.4 (80.0; 83.0) | 79.9 (78.4; 81.4) | 78.4 (77.0; 79.8) | 77.3 (75.9; 78.7) | 77.6 (76.2; 79.0) |
| **Overall, 0–64 years** | 77.6 (76.0; 79.2) | 76.2 (74.6; 77.8) | 74.9 (73.4; 76.5) | 74.4 (72.9; 75.9) | 74.5 (73.0; 76.0) |
| Female, 0–64 years | 62.8 (60.9; 64.8) | 61.0 (59.1; 62.9) | 59.9 (58.1; 61.8) | 59.1 (57.3; 61.0) | 59.3 (57.5; 61.2) |
| Male, 0–64 years | 92.0 (89.4; 94.6) | 90.9 (88.4; 93.5) | 89.5 (87.1; 92.0) | 89.2 (86.8; 91.6) | 89.2 (86.8; 91.7) |
| **Both sexes aged 0–17 years** | 0.6 (0.4; 1.0) | 0.7 (0.5; 1.1) | 0.8 (0.5; 1.2) | 0.7 (0.5; 1.1) | 0.5 (0.3; 0.8) |
| Females aged 0–17 years | 0.7 (0.4; 1.3) | 1.0 (0.6; 1.7) | 1.0 (0.6; 1.7) | 0.6 (0.3; 1.2) | 0.5 (0.2; 1.1) |
| Males aged 0–17 years | 0.5 (0.2; 1.1) | 0.5 (0.2; 1.0) | 0.6 (0.3; 1.2) | 0.8 (0.5; 1.4) | 0.5 (0.2; 1.0) |
| **Both sexes aged 18–64 years** | 98.7 (96.7; 100.8) | 96.9 (94.9; 98.9) | 95.3 (93.3; 97.3) | 94.6 (92.6; 96.5) | 94.8 (92.9; 96.7) |
| Females aged 18–64 years | 79.6 (77.1; 82.1) | 77.2 (74.9; 79.7) | 75.8 (73.5; 78.2) | 74.9 (72.6; 77.3) | 75.2 (72.9; 77.5) |
| Males aged 18–64 years | 117.5 (114.2; 120.8) | 116.1 (112.9; 119.4) | 114.3 (111.2; 117.5) | 113.8 (110.7; 116.9) | 113.9 (110.9; 117.0) |
| **Both sexes by age group (years)** | | | | | |
| 0–12 | 0.1 (0.0; 0.4) | 0.2 (0.1; 0.6) | 0.1 (0.0; 0.5) | 0.1 (0.0; 0.4) | 0 |
| 13–17 | 1.9 (1.1; 3.2) | 2.2 (1.4; 3.5) | 2.6 (1.6; 4.0) | 2.4 (1.5; 3.8) | 1.8 (1.1; 3.1) |
| 18–19 | 9.8 (6.6; 14.5) | 11.7 (8.4; 16.3) | 10.2 (7.2; 14.5) | 10.5 (7.5; 14.6) | 9.8 (7.0; 13.8) |
| 20–21 | 22.7 (17.8; 29.1) | 19.5 (15.2; 25.0) | 17.2 (13.2; 22.4) | 23.0 (18.4; 28.9) | 18.3 (14.3; 23.4) |
| 22–23 | 36.5 (30.3; 43.9) | 30.1 (24.8; 36.6) | 33.2 (27.5; 40.0) | 30.1 (24.8; 36.5) | 32.7 (27.2; 39.3) |
| 24–25 | 49.5 (42.3; 58.0) | 50.7 (43.8; 58.6) | 42.8 (36.6; 50.1) | 40.1 (34.2; 47.1) | 39.8 (33.8; 46.9) |
| 26–29 | 79.3 (73.0; 86.1) | 66.0 (60.4; 72.1) | 62.0 (56.8; 67.8) | 54.2 (49.5; 59.5) | 50.0 (45.5; 54.9) |
| 30–34 | 120.8 (113.8; 128.2) | 113.5 (106.9; 120.5) | 104.9 (98.7; 111.5) | 96.6 (90.8; 102.9) | 89.4 (83.9; 95.3) |
| 35–39 | 127.0 (119.4; 135.1) | 126.1 (118.7; 133.9) | 125.4 (118.3; 133.0) | 125.4 (118.5; 132.7) | 123.7 (117.0; 130.8) |
| 40–44 | 118.6 (112.0; 125.6) | 116.7 (110.0; 123.9) | 117.1 (110.2; 124.4) | 117.4 (110.5; 124.8) | 122.0 (114.9; 129.6) |
| 45–49 | 125.0 (119.1; 131.3) | 124.6 (118.7; 130.9) | 122.3 (116.4; 128.6) | 119.0 (113.1; 125.3) | 117.3 (111.2; 123.7) |
| 50–54 | 116.6 (110.7; 122.9) | 116.6 (110.8; 122.7) | 115.9 (110.3; 121.9) | 117.6 (112.0; 123.5) | 120.1 (114.5; 126.0) |
| 55–59 | 101.8 (95.8; 108.1) | 105.0 (99.1; 111.3) | 102.1 (96.4; 108.2) | 103.5 (97.8; 109.4) | 107.8 (102.1; 113.8) |
| 60–64 | 84.6 (78.8; 90.7) | 85.4 (79.8; 91.5) | 90.6 (84.8; 96.8) | 96.3 (90.4; 102.6) | 99.6 (93.6; 105.9) |

| | 2012 | 2013 | 2014 | 2015 | 2016 |
|---|---|---|---|---|---|
| **Female by age group (years)** | | | | | |
| 0–12 | 0.1 (0.0; 0.8) | 0.4 (0.1; 1.1) | 0.2 (0.1; 1.0) | 0.1 (0.0; 0.8) | 0 |
| 13–17 | 2.3 (1.1; 4.5) | 2.6 (1.4; 4.9) | 2.9 (1.6; 5.2) | 1.9 (0.9; 3.9) | 1.9 (0.9; 4.0) |
| 18–19 | 6.4 (3.2; 12.8) | 8.8 (5.1; 15.2) | 6.6 (3.5; 12.2) | 9.9 (6.1; 16.1) | 12.3 (8.0; 19.1) |
| 20–21 | 17.0 (11.4; 25.4) | 15.4 (10.3; 23.0) | 11.9 (7.6; 18.7) | 11.0 (6.9; 17.4) | 9.5 (5.8; 15.5) |
| 22–23 | 19.6 (13.8; 27.9) | 20.1 (14.4; 28.0) | 21.5 (15.5; 29.8) | 18.0 (12.6; 25.6) | 16.1 (11.1; 23.3) |
| 24–25 | 25.4 (18.7; 34.3) | 24.1 (18.0; 32.3) | 22.8 (16.9; 30.7) | 26.2 (19.9; 34.6) | 25.7 (19.3; 34.2) |
| 26–29 | 35.5 (29.9; 42.0) | 28.3 (23.5; 34.0) | 28.2 (23.5; 33.8) | 26.3 (21.9; 31.5) | 26.4 (22.0; 31.6) |
| 30–34 | 61.0 (54.4; 68.3) | 57.7 (51.5; 64.7) | 53.2 (47.3; 59.8) | 43.5 (38.3; 49.4) | 43.8 (38.6; 49.7) |
| 35–39 | 81.4 (73.5; 90.1) | 73.4 (66.1; 81.5) | 74.5 (67.3; 82.4) | 74.4 (67.4; 82.1) | 67.8 (61.2; 75.0) |
| 40–44 | 97.2 (89.5; 105.7) | 93.1 (85.4; 101.6) | 89.0 (81.3; 97.5) | 88.0 (80.3; 96.5) | 90.6 (82.6; 99.3) |
| 45–49 | 113.9 (106.5; 121.8) | 111.3 (104.1; 119.2) | 107.8 (100.6; 115.6) | 103.0 (95.8; 110.7) | 100.6 (93.3; 108.5) |
| 50–54 | 116.7 (108.9; 125.2) | 116.2 (108.6; 124.4) | 112.9 (105.5; 120.8) | 111.6 (104.4; 119.2) | 111.8 (104.6; 119.4) |
| 55–59 | 100.7 (92.9; 109.2) | 104.2 (96.4; 112.7) | 102.9 (95.3; 111.1) | 104.7 (97.3; 112.8) | 105.8 (98.4; 113.8) |
| 60–64 | 86.6 (78.9; 95.0) | 83.9 (76.6; 92.0) | 87.9 (80.4; 96.1) | 93.3 (85.6; 101.6) | 99.6 (91.7; 108.2) |
| **Male by age group (years)** | | | | | |
| 0–12 | 0.1 (0.0; 0.8) | 0 | 0 | 0 | 0 |
| 13–17 | 1.5 (0.6; 3.6) | 1.8 (0.8; 3.7) | 2.3 (1.2; 4.4) | 3.0 (1.7; 5.2) | 1.7 (0.8; 3.7) |
| 18–19 | 13.0 (8.1; 20.9) | 14.4 (9.5; 21.8) | 13.7 (9.0; 20.8) | 11.0 (7.0; 17.3) | 7.4 (4.3; 12.8) |
| 20–21 | 28.1 (20.6; 38.3) | 23.3 (16.9; 32.1) | 22.1 (16.0; 30.7) | 34.4 (26.6; 44.5) | 26.6 (20.0; 35.4) |
| 22–23 | 51.9 (41.8; 64.6) | 39.3 (31.0; 49.9) | 43.9 (35.0; 55.2) | 41.2 (32.7; 51.9) | 47.9 (38.7; 59.3) |
| 24–25 | 71.6 (59.5; 86.1) | 74.9 (63.3; 88.6) | 61.1 (50.9; 73.4) | 52.8 (43.4; 64.2) | 52.7 (43.2; 64.3) |
| 26–29 | 119.9 (109.1; 131.7) | 100.9 (91.2; 111.6) | 93.4 (84.4; 103.4) | 80.2 (72.1; 89.2) | 71.9 (64.4; 80.3) |
| 30–34 | 177.6 (165.6; 190.4) | 166.4 (155.1; 178.5) | 154.0 (143.5; 165.4) | 147.1 (137.0; 158.0) | 132.6 (123.2; 142.9) |
| 35–39 | 171.4 (158.6; 185.1) | 177.2 (164.6; 190.7) | 174.8 (162.8; 187.8) | 175.0 (163.3; 187.4) | 178.1 (166.6; 190.3) |
| 40–44 | 139.9 (129.3; 151.3) | 140.2 (129.3; 151.9) | 145.0 (133.8; 157.1) | 146.6 (135.3; 158.9) | 153.3 (141.7; 165.7) |
| 45–49 | 136.2 (127.1; 145.9) | 137.9 (128.7; 147.8) | 136.8 (127.6; 146.7) | 135.0 (125.7; 145.1) | 133.9 (124.4; 144.2) |
| 50–54 | 116.5 (107.8; 125.9) | 117.0 (108.5; 126.2) | 119.0 (110.5; 128.0) | 123.6 (115.2; 132.7) | 128.4 (119.9; 137.6) |
| 55–59 | 102.8 (94.0; 112.5) | 105.8 (97.0; 115.3) | 101.4 (93.0; 110.6) | 102.2 (93.9; 111.1) | 109.8 (101.3; 118.9) |
| 60–64 | 82.5 (74.2; 91.7) | 87.0 (78.6; 96.3) | 93.3 (84.7; 102.8) | 99.5 (90.7; 109.2) | 99.5 (90.7; 109.2) |

Estimates of the prevalence proportion of clozapine prescriptions are age- and sex-standardized to the population of Germany as of 31 December 2022.

**Table S1 (continued)**: Standardized prescription prevalence of clozapine by age and sex for each calendar year from 2012 to 2022 (per 100,000 persons; 95% confidence intervals in brackets)

| | 2017 | 2018 | 2019 | 2020 | 2021 | 2022 |
|---|---|---|---|---|---|---|
| **Total number of (database) population, 0–64 years, n** | 12,988,508 | 13,155,467 | 13,291,386 | 13,477,263 | 13,507,154 | 13,671,786 |
| Total number of (database) population, all age years, n | 16,412,811 | 16,649,554 | 16,853,824 | 17,115,837 | 17,219,027 | 17,469,599 |
| Overall, all age years | 75.9 (74.6; 77.3) | 75.0 (73.7; 76.4) | 74.1 (72.8; 75.4) | 72.5 (71.2; 73.8) | 71.7 (70.4; 73.0) | 69.4 (68.2; 70.6) |
| **Overall, 0–64 years** | 72.5 (71.1; 74.0) | 72.0 (70.5; 73.5) | 70.9 (69.4; 72.3) | 68.7 (67.3; 70.1) | 67.9 (66.5; 69.3) | 65.5 (64.2; 66.9) |
| Female, 0–64 years | 58.0 (56.2; 59.8) | 57.3 (55.6; 59.1) | 56.3 (54.5; 58.0) | 54.6 (52.9; 56.4) | 53.4 (51.7; 55.1) | 51.5 (49.8; 53.2) |
| Male, 0–64 years | 86.6 (84.3; 89.0) | 86.2 (84.0; 88.6) | 85.0 (82.8; 87.3) | 82.3 (80.1; 84.5) | 82.0 (79.8; 84.2) | 79.2 (77.0; 81.3) |
| **Both sexes aged 0–17 years** | 0.5 (0.3; 0.8) | 0.5 (0.3; 0.9) | 0.5 (0.3; 0.9) | 0.4 (0.2; 0.7) | 0.5 (0.3; 0.8) | 0.5 (0.3; 0.9) |
| Females aged 0–17 years | 0.6 (0.3; 1.2) | 0.6 (0.3; 1.2) | 0.7 (0.4; 1.3) | 0.5 (0.2; 1.1) | 0.6 (0.3; 1.3) | 0.4 (0.2; 1.0) |
| Males aged 0–17 years | 0.4 (0.1; 0.9) | 0.4 (0.2; 1.0) | 0.4 (0.2; 0.9) | 0.3 (0.1; 0.8) | 0.3 (0.1; 0.8) | 0.6 (0.3; 1.2) |
| **Both sexes aged 18–64 years** | 92.3 (90.4; 94.2) | 91.6 (89.8; 93.5) | 90.2 (88.3; 92.0) | 87.4 (85.6; 89.2) | 86.4 (84.7; 88.2) | 83.4 (81.6; 85.1) |
| Females aged 18–64 years | 73.5 (71.3; 75.8) | 72.6 (70.4; 74.9) | 71.3 (69.1; 73.5) | 69.2 (67.1; 71.5) | 67.7 (65.6; 69.9) | 65.3 (63.2; 67.4) |
| Males aged 18–64 years | 110.7 (107.7; 113.7) | 110.1 (107.2; 113.1) | 108.6 (105.7; 111.6) | 105.1 (102.3; 108.0) | 104.8 (102.0; 107.6) | 101.0 (98.3; 103.8) |
| **Both sexes by age group (years)** | | | | | | |
| 0–12 | 0 | 0 | 0 | 0.1 (0.0; 0.4) | 0.1 (0.0; 0.4) | 0.1 (0.0; 0.4) |
| 13–17 | 1.7 (1.0; 3.0) | 1.9 (1.1; 3.2) | 1.9 (1.1; 3.3) | 1.3 (0.7; 2.4) | 1.6 (0.9; 2.8) | 1.7 (1.0; 3.0) |
| 18–19 | 11.2 (8.1; 15.4) | 9.1 (6.4; 13.1) | 8.5 (5.9; 12.5) | 10.2 (7.1; 14.5) | 9.3 (6.5; 13.5) | 9.2 (6.3; 13.4) |
| 20–21 | 19.6 (15.6; 24.8) | 18.0 (14.2; 22.9) | 20.1 (16.0; 25.4) | 18.3 (14.3; 23.3) | 19.4 (15.2; 24.7) | 17.0 (13.1; 22.1) |
| 22–23 | 30.4 (25.2; 36.8) | 28.1 (23.2; 34.1) | 29.9 (24.8; 35.9) | 27.5 (22.7; 33.2) | 26.1 (21.4; 31.8) | 27.0 (22.2; 32.8) |
| 24–25 | 35.1 (29.6; 41.7) | 40.4 (34.4; 47.4) | 38.3 (32.6; 45.1) | 32.8 (27.6; 39.0) | 32.2 (27.1; 38.2) | 31.2 (26.2; 37.1) |
| 26–29 | 46.6 (42.3; 51.4) | 44.6 (40.4; 49.3) | 44.3 (40.1; 48.9) | 44.8 (40.5; 49.5) | 44.7 (40.4; 49.4) | 41.0 (37.0; 45.4) |
| 30–34 | 80.2 (75.0; 85.7) | 71.5 (66.8; 76.6) | 65.5 (61.0; 70.3) | 58.8 (54.7; 63.3) | 54.3 (50.4; 58.6) | 51.4 (47.5; 55.5) |
| 35–39 | 116.3 (109.9; 122.9) | 111.5 (105.5; 117.9) | 103.2 (97.5; 109.3) | 94.4 (89.0; 100.1) | 88.5 (83.3; 94.0) | 77.4 (72.6; 82.4) |
| 40–44 | 124.5 (117.5; 132.0) | 122.5 (115.6; 129.7) | 124.2 (117.4; 131.3) | 121.0 (114.5; 127.8) | 120.4 (114.0; 127.0) | 114.7 (108.7; 121.1) |
| 45–49 | 113.8 (107.6; 120.3) | 115.6 (109.1; 122.3) | 114.8 (108.3; 121.7) | 115.3 (108.7; 122.3) | 116.3 (109.6; 123.4) | 115.4 (108.8; 122.4) |
| 50–54 | 118.7 (113.1; 124.5) | 121.4 (115.7; 127.3) | 118.8 (113.1; 124.8) | 113.8 (108.1; 119.8) | 110.2 (104.5; 116.2) | 108.2 (102.4; 114.4) |
| 55–59 | 108.8 (103.3; 114.7) | 110.4 (104.9; 116.2) | 112.1 (106.7; 117.8) | 111.9 (106.5; 117.5) | 115.1 (109.7; 120.8) | 112.3 (107.0; 117.9) |
| 60–64 | 98.4 (92.5; 104.6) | 101.8 (95.9; 108.0) | 102.2 (96.5; 108.3) | 102.7 (97.1; 108.7) | 104.3 (98.7; 110.2) | 105.1 (99.6; 110.9) |

| | 2017 | 2018 | 2019 | 2020 | 2021 | 2022 |
|---|---|---|---|---|---|---|
| **Female by age group (years)** | | | | | | |
| 0–12 | 0 | 0 | 0 | 0.1 (0.0; 0.8) | 0.1 (0.0; 0.8) | 0 |
| 13–17 | 2.2 (1.1; 4.4) | 2.2 (1.1; 4.4) | 2.5 (1.3; 4.9) | 1.4 (0.6; 3.5) | 2.0 (1.0; 4.3) | 1.5 (0.6; 3.5) |
| 18–19 | 8.8 (5.2; 14.9) | 7.1 (3.9; 12.8) | 10.5 (6.5; 17.2) | 11.0 (6.7; 18.0) | 11.1 (6.8; 18.1) | 13.4 (8.5; 21.0) |
| 20–21 | 13.1 (8.7; 19.8) | 13.1 (8.7; 19.7) | 14.0 (9.4; 20.9) | 14.9 (10.1; 22.1) | 14.0 (9.3; 21.1) | 12.6 (8.1; 19.5) |
| 22–23 | 13.7 (9.2; 20.5) | 15.0 (10.3; 21.9) | 21.0 (15.3; 28.7) | 18.7 (13.4; 26.1) | 15.1 (10.4; 22.0) | 17.5 (12.3; 24.9) |
| 24–25 | 21.2 (15.5; 29.0) | 20.4 (14.9; 28.1) | 18.2 (13.0; 25.5) | 18.0 (13.0; 25.1) | 22.4 (16.7; 30.1) | 19.3 (14.1; 26.6) |
| 26–29 | 25.5 (21.3; 30.6) | 25.1 (20.8; 30.2) | 24.8 (20.6; 29.9) | 25.3 (21.0; 30.5) | 24.5 (20.3; 29.6) | 24.2 (20.0; 29.3) |
| 30–34 | 40.8 (35.8; 46.4) | 38.7 (34.0; 44.1) | 35.4 (31.0; 40.6) | 33.5 (29.2; 38.3) | 30.5 (26.5; 35.2) | 29.8 (25.8; 34.4) |
| 35–39 | 64.9 (58.6; 71.9) | 62.8 (56.7; 69.6) | 54.9 (49.3; 61.2) | 46.3 (41.2; 52.1) | 47.4 (42.2; 53.2) | 42.8 (38.0; 48.3) |
| 40–44 | 83.7 (76.1; 92.1) | 78.9 (71.7; 87.0) | 78.2 (71.1; 86.1) | 78.3 (71.3; 86.0) | 71.7 (65.1; 78.9) | 65.5 (59.3; 72.3) |
| 45–49 | 100.2 (92.7; 108.4) | 98.3 (90.6; 106.7) | 95.8 (88.0; 104.3) | 93.8 (85.9; 102.4) | 93.5 (85.5; 102.1) | 83.4 (75.9; 91.5) |
| 50–54 | 112.2 (105.0; 119.8) | 111.0 (103.9; 118.7) | 107.9 (100.8; 115.6) | 101.1 (94.1; 108.7) | 97.6 (90.5; 105.3) | 99.6 (92.2; 107.5) |
| 55–59 | 106.5 (99.2; 114.4) | 108.2 (101.0; 116.0) | 110.1 (102.9; 117.7) | 107.8 (100.8; 115.3) | 108.3 (101.3; 115.8) | 104.9 (98.1; 112.3) |
| 60–64 | 98.5 (90.8; 106.9) | 100.5 (92.8; 108.8) | 100.6 (93.0; 108.8) | 103.8 (96.3; 111.9) | 101.8 (94.5; 109.6) | 102.1 (94.9; 109.8) |
| **Male by age group (years)** | | | | | | |
| 0–12 | 0 | 0 | 0 | 0 | 0 | 0.1 (0.0; 0.7) |
| 13–17 | 1.3 (0.5; 3.1) | 1.6 (0.7; 3.5) | 1.4 (0.6; 3.3) | 1.1 (0.4; 3.0) | 1.1 (0.4; 2.9) | 1.9 (0.9; 4.1) |
| 18–19 | 13.4 (8.9; 20.2) | 11.1 (7.1; 17.4) | 6.7 (3.7; 12.0) | 9.4 (5.7; 15.6) | 7.7 (4.4; 13.5) | 5.2 (2.6; 10.4) |
| 20–21 | 25.7 (19.4; 34.1) | 22.7 (16.8; 30.6) | 25.9 (19.5; 34.3) | 21.4 (15.6; 29.3) | 24.4 (18.1; 32.9) | 21.2 (15.3; 29.4) |
| 22–23 | 45.7 (36.9; 56.7) | 40.1 (32.0; 50.2) | 38.0 (30.3; 47.6) | 35.5 (28.1; 44.8) | 36.1 (28.6; 45.7) | 35.6 (28.1; 45.2) |
| 24–25 | 47.8 (38.9; 58.8) | 58.6 (48.7; 70.5) | 56.7 (47.1; 68.4) | 46.2 (37.8; 56.6) | 41.1 (33.3; 50.8) | 42.0 (34.1; 51.7) |
| 26–29 | 66.2 (59.1; 74.2) | 62.7 (55.8; 70.6) | 62.3 (55.4; 70.1) | 62.8 (55.8; 70.6) | 63.4 (56.4; 71.3) | 56.5 (50.0; 63.9) |
| 30–34 | 117.5 (108.8; 127.0) | 102.7 (94.7; 111.4) | 94.0 (86.5; 102.1) | 82.9 (76.0; 90.4) | 76.9 (70.3; 84.1) | 71.8 (65.5; 78.8) |
| 35–39 | 166.1 (155.4; 177.5) | 158.9 (148.7; 169.8) | 150.1 (140.4; 160.5) | 141.0 (131.7; 150.9) | 128.4 (119.6; 137.8) | 110.9 (102.8; 119.5) |
| 40–44 | 165.1 (153.4; 177.7) | 165.7 (154.3; 178.0) | 169.8 (158.5; 181.9) | 163.4 (152.7; 174.8) | 168.7 (158.1; 180.1) | 163.7 (153.5; 174.6) |
| 45–49 | 127.3 (117.8; 137.6) | 132.8 (122.7; 143.7) | 133.7 (123.5; 144.9) | 136.7 (126.3; 148.1) | 139.1 (128.6; 150.5) | 147.4 (136.8; 158.9) |
| 50–54 | 125.2 (116.8; 134.2) | 131.7 (123.0; 140.9) | 129.7 (121.0; 139.0) | 126.5 (117.7; 135.8) | 122.6 (113.8; 132.1) | 116.8 (108.0; 126.3) |
| 55–59 | 111.1 (102.9; 120.1) | 112.5 (104.4; 121.3) | 114.1 (106.1; 122.8) | 115.9 (107.9; 124.6) | 122.0 (113.8; 130.7) | 119.6 (111.6; 128.2) |
| 60–64 | 98.3 (89.6; 107.7) | 103.2 (94.5; 112.7) | 103.9 (95.4; 113.3) | 101.7 (93.4; 110.7) | 106.9 (98.6; 115.9) | 108.2 (100.0; 117.0) |

Estimates of the prevalence proportion of clozapine prescriptions are age- and sex-standardized to the population of Germany as of 31 December 2022.

**Table S2:** Standardized prescription incidence of clozapine by age and sex for each calendar year from 2012 to 2022 (per 100,000 persons; 95% confidence intervals in brackets)

| | **2012** | **2013** | **2014** | **2015** | **2016** |
|---|---|---|---|---|---|
| **Total number of (database) population, 0–64 years, n** | 10,161,228 | 11,308,968 | 11,754,464 | 11,974,203 | 12,071,904 |
| **Overall, 0–64 years** | 7.1 (6.6; 7.7) | 6.6 (6.1; 7.1) | 6.1 (5.6; 6.5) | 6.4 (5.9; 6.9) | 6.1 (5.7; 6.6) |
| Female, 0–64 years | 6.0 (5.4; 6.7) | 5.4 (4.8; 6.0) | 4.7 (4.2; 5.3) | 4.9 (4.4; 5.5) | 4.9 (4.4; 5.5) |
| Male, 0–64 years | 8.2 (7.4; 9.1) | 7.8 (7.0; 8.6) | 7.4 (6.7; 8.2) | 7.8 (7.1; 8.5) | 7.3 (6.6; 8.0) |
| **Both sexes aged 0–17 years** | 0.4 (0.2; 0.7) | 0.4 (0.2; 0.8) | 0.4 (0.2; 0.8) | 0.4 (0.3; 0.8) | 0.2 (0.1; 0.4) |
| Females aged 0–17 years | 0.4 (0.2; 1.1) | 0.4 (0.2; 1.0) | 0.5 (0.2; 1.1) | 0.3 (0.1; 0.8) | 0.2 (0.1; 0.7) |
| Males aged 0–17 years | 0.3 (0.1; 0.9) | 0.4 (0.2; 1.0) | 0.4 (0.2; 0.9) | 0.6 (0.3; 1.1) | 0.1 (0.0; 0.6) |
| **Both sexes aged 18–64 years** | 9.0 (8.3; 9.7) | 8.3 (7.7; 8.9) | 7.6 (7.1; 8.2) | 8.0 (7.4; 8.6) | 7.7 (7.2; 8.3) |
| Females aged 18–64 years | 7.5 (6.7; 8.4) | 6.7 (6.0; 7.5) | 5.8 (5.2; 6.5) | 6.2 (5.5; 6.9) | 6.2 (5.6; 7.0) |
| Males aged 18–64 years | 10.4 (9.4; 11.5) | 9.8 (8.9; 10.8) | 9.4 (8.5; 10.4) | 9.8 (8.9; 10.7) | 9.2 (8.4; 10.2) |
| **Both sexes by age group (years)** | | | | | |
| 0–12 | 0.1 (0.0; 0.4) | 0.1 (0.0; 0.5) | 0.1 (0.0; 0.5) | 0.1 (0.0; 0.4) | 0 |
| 13–17 | 1.2 (0.6; 2.5) | 1.2 (0.6; 2.4) | 1.2 (0.6; 2.3) | 1.5 (0.8; 2.7) | 0.7 (0.3; 1.6) |
| 18–19 | 5.4 (3.0; 9.8) | 6.2 (3.7; 10.2) | 5.5 (3.4; 8.9) | 4.9 (2.9; 8.0) | 3.5 (1.9; 6.3) |
| 20–21 | 9.2 (5.9; 14.2) | 10.2 (7.0; 15.1) | 5.5 (3.4; 9.0) | 8.8 (6.0; 12.9) | 6.1 (3.9; 9.5) |
| 22–23 | 9.3 (6.2; 14.0) | 8.4 (5.6; 12.5) | 9.6 (6.7; 13.9) | 10.3 (7.3; 14.6) | 11.3 (8.1; 15.6) |
| 24–25 | 9.4 (6.3; 14.2) | 11.2 (7.9; 15.9) | 9.9 (7.0; 13.9) | 7.2 (4.8; 10.7) | 8.3 (5.7; 12.1) |
| 26–29 | 11.6 (9.2; 14.7) | 7.7 (5.8; 10.1) | 9.2 (7.2; 11.7) | 9.0 (7.1; 11.4) | 8.3 (6.5; 10.6) |
| 30–34 | 12.8 (10.5; 15.5) | 10.9 (8.9; 13.3) | 10.6 (8.7; 12.9) | 10.0 (8.2; 12.3) | 11.1 (9.2; 13.4) |
| 35–39 | 8.6 (6.6; 11.0) | 9.4 (7.5; 11.8) | 10.4 (8.5; 12.8) | 10.9 (8.9; 13.3) | 9.0 (7.3; 11.2) |
| 40–44 | 7.9 (6.2; 10.1) | 9.0 (7.2; 11.2) | 6.7 (5.2; 8.7) | 7.7 (6.1; 9.9) | 7.1 (5.6; 9.2) |
| 45–49 | 8.4 (6.9; 10.3) | 7.2 (5.9; 8.9) | 6.7 (5.4; 8.3) | 6.6 (5.3; 8.2) | 6.6 (5.3; 8.4) |
| 50–54 | 7.3 (5.8; 9.1) | 6.2 (4.9; 7.8) | 5.7 (4.5; 7.2) | 6.7 (5.5; 8.3) | 6.2 (5.0; 7.7) |
| 55–59 | 7.6 (6.1; 9.6) | 7.3 (5.8; 9.1) | 5.2 (4.0; 6.7) | 6.7 (5.3; 8.3) | 6.1 (4.8; 7.6) |
| 60–64 | 9.3 (7.4; 11.6) | 7.7 (6.2; 9.8) | 7.4 (5.9; 9.4) | 7.2 (5.7; 9.1) | 8.3 (6.7; 10.3) |

Estimates of the incidence proportion of clozapine prescriptions are age- and sex-standardized to the population of Germany as of 31 December 2022.

**Table S2 (continued)**: Standardized prescription incidence of clozapine by age and sex for each calendar year from 2012 to 2022 (per 100,000 persons; 95% confidence intervals in brackets)

| | **2017** | **2018** | **2019** | **2020** | **2021** | **2022** |
|---|---|---|---|---|---|---|
| **Total number of (database) population, 0–64 years, n** | 12,250,701 | 12,566,794 | 12,732,135 | 12,900,409 | 13,019,346 | 13,077,244 |
| **Overall, 0–64 years** | 5.6 (5.2; 6.1) | 5.5 (5.1; 6.0) | 5.4 (5.1; 5.9) | 4.7 (4.3; 5.1) | 4.7 (4.3; 5.1) | 4.2 (3.8; 4.5) |
| Female, 0–64 years | 4.6 (4.1; 5.2) | 4.3 (3.8; 4.8) | 4.4 (3.9; 4.9) | 3.5 (3.1; 4.0) | 3.6 (3.2; 4.1) | 3.2 (2.8; 3.6) |
| Male, 0–64 years | 6.6 (5.9; 7.3) | 6.8 (6.1; 7.5) | 6.5 (5.9; 7.2) | 5.8 (5.3; 6.5) | 5.8 (5.2; 6.4) | 5.1 (4.6; 5.7) |
| **Both sexes aged 0–17 years** | 0.3 (0.2; 0.7) | 0.2 (0.1; 0.5) | 0.3 (0.2; 0.6) | 0.2 (0.1; 0.5) | 0.3 (0.1; 0.6) | 0.4 (0.2; 0.7) |
| Females aged 0–17 years | 0.5 (0.2; 1.1) | 0.2 (0.1; 0.7) | 0.4 (0.2; 1.0) | 0.4 (0.2; 1.0) | 0.4 (0.2; 1.0) | 0.2 (0.1; 0.8) |
| Males aged 0–17 years | 0.2 (0.1; 0.7) | 0.2 (0.1; 0.7) | 0.2 (0.1; 0.7) | 0.1 (0.0; 0.6) | 0.2 (0.0; 0.6) | 0.5 (0.2; 1.0) |
| **Both sexes aged 18–64 years** | 7.1 (6.6; 7.6) | 7.0 (6.5; 7.5) | 6.9 (6.4; 7.4) | 5.9 (5.5; 6.4) | 5.9 (5.4; 6.4) | 5.2 (4.8; 5.7) |
| Females aged 18–64 years | 5.8 (5.1; 6.5) | 5.3 (4.8; 6.0) | 5.4 (4.8; 6.1) | 4.3 (3.8; 4.9) | 4.4 (3.9; 5.0) | 4.0 (3.5; 4.6) |
| Males aged 18–64 years | 8.3 (7.5; 9.2) | 8.6 (7.8; 9.5) | 8.2 (7.5; 9.1) | 7.4 (6.7; 8.2) | 7.3 (6.6; 8.1) | 6.4 (5.7; 7.1) |
| **Both sexes by age group (years)** | | | | | | |
| 0–12 | 0 | 0 | 0 | 0.1 (0.0; 0.4) | 0 | 0.1 (0.0; 0.4) |
| 13–17 | 1.3 (0.7; 2.4) | 0.8 (0.4; 1.9) | 1.2 (0.6; 2.3) | 0.7 (0.3; 1.7) | 1.0 (0.5; 2.1) | 1.2 (0.6; 2.4) |
| 18–19 | 5.2 (3.2; 8.5) | 4.2 (2.5; 7.3) | 4.3 (2.5; 7.5) | 4.9 (2.9; 8.2) | 4.6 (2.7; 7.9) | 4.3 (2.5; 7.6) |
| 20–21 | 8.9 (6.2; 12.8) | 7.1 (4.8; 10.6) | 7.0 (4.6; 10.5) | 7.1 (4.7; 10.7) | 7.6 (5.1; 11.4) | 4.6 (2.7; 7.8) |
| 22–23 | 8.1 (5.5; 11.8) | 7.0 (4.7; 10.4) | 9.8 (7.0; 13.6) | 7.2 (4.9; 10.6) | 5.7 (3.7; 8.9) | 7.9 (5.4; 11.5) |
| 24–25 | 8.2 (5.6; 12.0) | 8.9 (6.2; 12.7) | 8.4 (5.8; 12.1) | 5.9 (3.8; 9.0) | 8.3 (5.8; 11.8) | 8.1 (5.7; 11.6) |
| 26–29 | 7.0 (5.4; 9.1) | 8.9 (7.0; 11.2) | 7.9 (6.1; 10.1) | 8.4 (6.6; 10.7) | 6.9 (5.3; 9.0) | 5.5 (4.1; 7.4) |
| 30–34 | 8.1 (6.5; 10.0) | 8.4 (6.9; 10.4) | 7.3 (5.9; 9.1) | 7.0 (5.7; 8.7) | 5.0 (3.9; 6.5) | 5.0 (3.9; 6.5) |
| 35–39 | 9.6 (7.9; 11.7) | 7.2 (5.7; 9.0) | 8.3 (6.7; 10.2) | 5.8 (4.6; 7.4) | 6.2 (4.9; 7.8) | 5.6 (4.4; 7.2) |
| 40–44 | 6.7 (5.2; 8.7) | 6.7 (5.3; 8.7) | 6.2 (4.8; 7.9) | 5.8 (4.5; 7.5) | 5.6 (4.4; 7.3) | 4.9 (3.7; 6.4) |
| 45–49 | 6.2 (4.9; 8.0) | 6.9 (5.5; 8.7) | 5.8 (4.5; 7.6) | 5.0 (3.8; 6.7) | 4.6 (3.4; 6.3) | 3.4 (2.4; 4.8) |
| 50–54 | 5.3 (4.2; 6.6) | 6.2 (5.0; 7.7) | 6.0 (4.8; 7.5) | 4.7 (3.7; 6.1) | 4.7 (3.6; 6.1) | 4.9 (3.8; 6.3) |
| 55–59 | 6.2 (5.0; 7.8) | 6.3 (5.1; 7.8) | 5.6 (4.5; 7.0) | 4.3 (3.3; 5.5) | 5.8 (4.6; 7.2) | 4.6 (3.6; 5.9) |
| 60–64 | 7.0 (5.5; 8.9) | 6.1 (4.7; 7.7) | 7.4 (5.9; 9.2) | 6.7 (5.4; 8.4) | 7.4 (6.0; 9.2) | 6.1 (4.9; 7.7) |

Estimates of the incidence proportion of clozapine prescriptions are age- and sex-standardized to the population of Germany as of 31 December 2022.

**Table S3:** Standardized prescription prevalence of clozapine (age 0–64 years) by regional characteristics for each calendar year from 2012 to 2022 (per 100,000 persons; 95% confidence intervals in brackets)

| | **2012** | **2013** | **2014** | **2015** | **2016** |
|---|---|---|---|---|---|
| Total number of (database) population, n | 11,650,141 | 12,130,060 | 12,372,493 | 12,658,252 | 12,733,756 |
| **Overall** | 77.6 (76.0; 79.3) | 76.2 (74.6; 77.8) | 74.9 (73.4; 76.5) | 74.4 (72.8; 75.9) | 74.5 (73.0; 76.0) |
| **Type of district according to settlement structure, i.e., urbanicity** | | | | | |
| Large urban city | 94.7 (91.6; 98.0) | 91.9 (88.9; 95.0) | 90.3 (87.4; 93.4) | 89.6 (86.7; 92.6) | 90.2 (87.3; 93.2) |
| Urban district | 74.6 (72.1; 77.3) | 73.0 (70.6; 75.6) | 71.7 (69.3; 74.2) | 70.1 (67.8; 72.6) | 69.9 (67.6; 72.3) |
| Rural district (with densification tendencies) | 64.6 (60.8; 68.6) | 65.4 (61.6; 69.4) | 64.8 (61.1; 68.7) | 66.5 (62.8; 70.4) | 66.5 (62.7; 70.4) |
| Sparsely populated rural district | 64.1 (60.1; 68.4) | 64.8 (60.8; 69.1) | 63.7 (59.7; 67.9) | 63.4 (59.5; 67.6) | 63.3 (59.5; 67.5) |
| **German Index of Socioeconomic Deprivation (2018)** | | | | | |
| 1st quintile (least deprived) | 88.7 (85.3; 92.3) | 85.6 (82.3; 89.1) | 81.9 (78.7; 85.3) | 81.0 (77.8; 84.2) | 81.4 (78.2; 84.6) |
| 2nd to 4th quintile | 74.7 (72.7; 76.8) | 73.8 (71.8; 75.8) | 73.4 (71.5; 75.4) | 72.7 (70.8; 74.7) | 73.0 (71.1; 75.0) |
| 5th quintile (most deprived) | 72.6 (68.5; 77.0) | 72.0 (68.0; 76.3) | 71.3 (67.3; 75.6) | 71.5 (67.5; 75.7) | 70.2 (66.3; 74.4) |

Estimates of the prevalence proportion of clozapine prescriptions are age- and sex-standardized to the population of Germany as of 31 December 2022.

**Table S3 (continued)**: Standardized prescription prevalence of clozapine (age 0–64 years) by regional characteristics for each calendar year from 2012 to 2022 (per 100,000 persons; 95% confidence intervals in brackets)

| | **2017** | **2018** | **2019** | **2020** | **2021** | **2022** |
|---|---|---|---|---|---|---|
| Total number of (database) population, n | 12,987,202 | 13,155,155 | 13,291,178 | 13,477,138 | 13,507,036 | 13,671,677 |
| **Overall** | 72.5 (71.1; 74.0) | 72.0 (70.5; 73.5) | 70.9 (69.4; 72.3) | 68.6 (67.3; 70.1) | 67.9 (66.5; 69.3) | 65.5 (64.2; 66.9) |
| **Type of district according to settlement structure, i.e., urbanicity** | | | | | | |
| Large urban city | 85.9 (83.1; 88.8) | 84.3 (81.6; 87.1) | 81.5 (78.9; 84.3) | 78.0 (75.4; 80.6) | 76.3 (73.8; 78.9) | 73.1 (70.7; 75.7) |
| Urban district | 68.4 (66.1; 70.7) | 68.1 (65.8; 70.4) | 67.3 (65.1; 69.6) | 65.5 (63.3; 67.7) | 64.6 (62.4; 66.8) | 63.0 (60.9; 65.2) |
| Rural district (with densification tendencies) | 67.7 (64.0; 71.6) | 67.3 (63.6; 71.2) | 67.0 (63.3; 70.9) | 65.5 (61.9; 69.3) | 65.3 (61.7; 69.1) | 62.4 (58.9; 66.1) |
| Sparsely populated rural district | 62.0 (58.2; 66.1) | 63.2 (59.3; 67.3) | 64.3 (60.5; 68.5) | 63.9 (60.1; 68.0) | 65.6 (61.7; 69.8) | 63.0 (59.2; 67.1) |
| **German Index of Socioeconomic Deprivation (2018)** | | | | | | |
| 1st quintile (least deprived) | 78.9 (75.9; 82.1) | 77.7 (74.8; 80.8) | 75.6 (72.7; 78.6) | 73.4 (70.6; 76.3) | 71.4 (68.7; 74.3) | 69.2 (66.5; 72.0) |
| 2nd to 4th quintile | 71.3 (69.5; 73.2) | 70.8 (69.0; 72.7) | 69.7 (67.9; 71.6) | 67.4 (65.6; 69.2) | 66.9 (65.2; 68.7) | 64.2 (62.5; 65.9) |
| 5th quintile (most deprived) | 67.7 (63.9; 71.8) | 68.1 (64.3; 72.2) | 68.6 (64.8; 72.7) | 66.8 (63.0; 70.9) | 67.2 (63.4; 71.3) | 66.2 (62.5; 70.2) |

Estimates of the prevalence proportion of clozapine prescriptions are age- and sex-standardized to the population of Germany as of 31 December 2022.

**Table S4:** Prescription prevalence of clozapine (age 0–64 years) by district among the 202 districts with a database population of ≥20,000 persons in 2022 (per 100,000 persons), in ascending order of the standardized prevalence

| District ID | Name of district | Urbanicity | Federal state | Database population, n | Clozapine users, n | Crude prevalence | Standardized prevalence | 95% CI |
|---|---|---|---|---|---|---|---|---|
| 3458 | Oldenburg | Rural district (with densification tendencies) | Niedersachsen | 32512 | 2 | 6.2 | 5.4 | (1.4; 21.7) |
| 9775 | Neu-Ulm | Urban district | Bayern | 26560 | 5 | 18.8 | 17.9 | (7.4; 43.0) |
| 3457 | Leer | Rural district (with densification tendencies) | Niedersachsen | 33446 | 7 | 20.9 | 19.9 | (9.5; 41.7) |
| 12065 | Oberhavel | Sparsely populated rural district | Brandenburg | 43065 | 10 | 23.2 | 21.5 | (11.5; 40.2) |
| 3461 | Wesermarsch | Sparsely populated rural district | Niedersachsen | 23382 | 6 | 25.7 | 22.6 | (10.1; 51.0) |
| 15003 | Magdeburg, Stadt | Large urban city | Sachsen-Anhalt | 30246 | 7 | 23.1 | 23.4 | (11.0; 49.7) |
| 7138 | Neuwied | Urban district | Rheinland-Pfalz | 35915 | 9 | 25.1 | 24.1 | (12.5; 46.5) |
| 9572 | Erlangen-Höchstadt | Urban district | Bayern | 24326 | 6 | 24.7 | 26.5 | (11.9; 59.3) |
| 9574 | Nürnberger Land | Urban district | Bayern | 25689 | 7 | 27.2 | 27.7 | (13.1; 58.7) |
| 12061 | Dahme-Spreewald | Sparsely populated rural district | Brandenburg | 33317 | 9 | 27.0 | 28.7 | (14.5; 56.5) |
| 9772 | Augsburg | Urban district | Bayern | 38279 | 11 | 28.7 | 29.7 | (16.5; 53.6) |
| 12069 | Potsdam-Mittelmark | Rural district (with densification tendencies) | Brandenburg | 42706 | 13 | 30.4 | 29.8 | (17.1; 52.0) |
| 5978 | Unna | Urban district | Nordrhein-Westfalen | 60526 | 19 | 31.4 | 29.9 | (19.0; 47.1) |
| 3451 | Ammerland | Urban district | Niedersachsen | 27139 | 8 | 29.5 | 31.1 | (15.5; 62.5) |
| 3453 | Cloppenburg | Rural district (with densification tendencies) | Niedersachsen | 41170 | 13 | 31.6 | 31.5 | (18.3; 54.2) |
| 7133 | Bad Kreuznach | Rural district (with densification tendencies) | Rheinland-Pfalz | 24822 | 9 | 36.3 | 31.6 | (16.3; 61.4) |
| 7143 | Westerwaldkreis | Urban district | Rheinland-Pfalz | 36772 | 11 | 29.9 | 31.6 | (17.4; 57.4) |
| 9175 | Ebersberg | Urban district | Bayern | 27644 | 9 | 32.6 | 31.7 | (16.3; 61.3) |
| 3361 | Verden | Rural district (with densification tendencies) | Niedersachsen | 33193 | 12 | 36.2 | 33.1 | (18.7; 58.7) |
| 12054 | Potsdam, Stadt | Large urban city | Brandenburg | 44103 | 14 | 31.7 | 33.2 | (19.4; 56.7) |
| 3459 | Osnabrück | Rural district (with densification tendencies) | Niedersachsen | 60392 | 19 | 31.5 | 33.5 | (21.3; 52.5) |
| 6633 | Kassel | Urban district | Hessen | 45727 | 15 | 32.8 | 34.6 | (20.8; 57.5) |
| 1062 | Stormarn | Urban district | Schleswig-Holstein | 76313 | 25 | 32.8 | 34.8 | (23.4; 51.8) |

| District ID | Name of district | Urbanicity | Federal state | Database population, n | Clozapine users, n | Crude prevalence | Standardized prevalence | 95% CI |
|---|---|---|---|---|---|---|---|---|
| 5554 | Borken | Urban district | Nordrhein-Westfalen | 71615 | 26 | 36.3 | 35.5 | (24.1; 52.2) |
| 13072 | Landkreis Rostock | Sparsely populated rural district | Mecklenburg-Vorpommern | 48661 | 18 | 37.0 | 35.5 | (22.2; 56.8) |
| 5378 | Rheinisch-Berg. Kr. | Urban district | Nordrhein-Westfalen | 47799 | 17 | 35.6 | 35.9 | (22.3; 57.9) |
| 3356 | Osterholz | Urban district | Niedersachsen | 29821 | 10 | 33.5 | 36.0 | (19.2; 67.7) |
| 5170 | Wesel | Urban district | Nordrhein-Westfalen | 70815 | 26 | 36.7 | 36.5 | (24.8; 53.8) |
| 5913 | Dortmund, Stadt | Large urban city | Nordrhein-Westfalen | 103180 | 38 | 36.8 | 36.7 | (26.6; 50.5) |
| 9671 | Aschaffenburg | Urban district | Bayern | 29424 | 11 | 37.4 | 36.8 | (20.3; 66.8) |
| 3403 | Oldenburg, Stadt | Large urban city | Niedersachsen | 38902 | 15 | 38.6 | 36.9 | (22.2; 61.3) |
| 5954 | Ennepe-Ruhr-Kreis | Urban district | Nordrhein-Westfalen | 59096 | 23 | 38.9 | 37.0 | (24.5; 55.8) |
| 7132 | Altenkir. (Westerw.) | Urban district | Rheinland-Pfalz | 21020 | 8 | 38.1 | 37.2 | (18.5; 74.8) |
| 3455 | Friesland | Urban district | Niedersachsen | 21321 | 8 | 37.5 | 37.3 | (18.4; 75.7) |
| 6631 | Fulda | Rural district (with densification tendencies) | Hessen | 46700 | 17 | 36.4 | 37.3 | (23.2; 60.2) |
| 5374 | Oberbergischer Kreis | Urban district | Nordrhein-Westfalen | 42801 | 16 | 37.4 | 37.9 | (23.2; 62.0) |
| 13075 | Vorp.-Greifswald | Sparsely populated rural district | Mecklenburg-Vorpommern | 41079 | 16 | 38.9 | 38.5 | (23.2; 63.8) |
| 5117 | Mülheim a.d.R., St. | Large urban city | Nordrhein-Westfalen | 29307 | 11 | 37.5 | 38.6 | (21.4; 69.9) |
| 3353 | Harburg | Urban district | Niedersachsen | 66650 | 26 | 39.0 | 38.9 | (26.4; 57.2) |
| 9188 | Starnberg | Urban district | Bayern | 20495 | 7 | 34.2 | 38.9 | (18.4; 82.1) |
| 3352 | Cuxhaven | Sparsely populated rural district | Niedersachsen | 42360 | 18 | 42.5 | 39.0 | (24.4; 62.2) |
| 3359 | Stade | Rural district (with densification tendencies) | Niedersachsen | 45048 | 17 | 37.7 | 39.2 | (24.4; 63.3) |
| 6434 | Hochtaunuskreis | Urban district | Hessen | 55186 | 22 | 39.9 | 39.4 | (25.8; 60.3) |
| 1054 | Nordfriesland | Sparsely populated rural district | Schleswig-Holstein | 28840 | 11 | 38.1 | 39.8 | (21.6; 73.5) |
| 9178 | Freising | Urban district | Bayern | 29361 | 12 | 40.9 | 39.8 | (22.6; 70.3) |
| 5316 | Leverkusen, Stadt | Large urban city | Nordrhein-Westfalen | 20800 | 8 | 38.5 | 40.1 | (19.9; 80.5) |
| 3452 | Aurich | Rural district (with densification tendencies) | Niedersachsen | 26792 | 11 | 41.1 | 40.6 | (22.5; 73.4) |
| 13074 | Nordwestmecklenburg | Rural district (with densification tendencies) | Mecklenburg-Vorpommern | 29767 | 11 | 37.0 | 40.6 | (22.2; 74.3) |
| 6433 | Groß-Gerau | Urban district | Hessen | 48872 | 20 | 40.9 | 41.5 | (26.7; 64.3) |
| 9563 | Fürth, Stadt | Large urban city | Bayern | 20097 | 9 | 44.8 | 42.0 | (21.8; 81.1) |

| District ID | Name of district | Urbanicity | Federal state | Database population, n | Clozapine users, n | Crude prevalence | Standardized prevalence | 95% CI |
|---|---|---|---|---|---|---|---|---|
| 13076 | Ludwigslust-Parchim | Sparsely populated rural district | Mecklenburg-Vorpommern | 36722 | 16 | 43.6 | 42.3 | (25.6; 70.0) |
| 9179 | Fürstenfeldbruck | Urban district | Bayern | 38996 | 16 | 41.0 | 43.1 | (26.3; 70.6) |
| 8426 | Biberach | Rural district (with densification tendencies) | Baden-Württemberg | 21710 | 10 | 46.1 | 43.3 | (23.2; 80.9) |
| 12064 | Märkisch-Oderland | Sparsely populated rural district | Brandenburg | 38538 | 17 | 44.1 | 43.6 | (26.7; 71.1) |
| 5362 | Rhein-Erft-Kreis | Urban district | Nordrhein-Westfalen | 87060 | 39 | 44.8 | 43.9 | (32.0; 60.1) |
| 12067 | Oder-Spree | Sparsely populated rural district | Brandenburg | 29208 | 14 | 47.9 | 43.9 | (25.9; 74.3) |
| 12063 | Havelland | Sparsely populated rural district | Brandenburg | 31988 | 13 | 40.6 | 44.0 | (25.0; 77.4) |
| 3151 | Gifhorn | Sparsely populated rural district | Niedersachsen | 20034 | 8 | 39.9 | 44.4 | (21.9; 89.8) |
| 13073 | Vorpommern-Rügen | Sparsely populated rural district | Mecklenburg-Vorpommern | 36825 | 17 | 46.2 | 44.5 | (27.6; 71.8) |
| 5119 | Oberhausen, Stadt | Large urban city | Nordrhein-Westfalen | 26335 | 12 | 45.6 | 44.6 | (25.3; 78.5) |
| 5758 | Herford | Urban district | Nordrhein-Westfalen | 28539 | 13 | 45.6 | 44.6 | (25.8; 77.0) |
| 3454 | Emsland | Sparsely populated rural district | Niedersachsen | 47688 | 22 | 46.1 | 44.8 | (29.5; 68.0) |
| 9174 | Dachau | Urban district | Bayern | 21368 | 9 | 42.1 | 45.4 | (23.5; 87.9) |
| 3251 | Diepholz | Rural district (with densification tendencies) | Niedersachsen | 43850 | 20 | 45.6 | 45.5 | (29.2; 70.9) |
| 6438 | Offenbach | Urban district | Hessen | 70570 | 33 | 46.8 | 45.8 | (32.6; 64.6) |
| 12072 | Teltow-Fläming | Sparsely populated rural district | Brandenburg | 31270 | 10 | 32.0 | 45.9 | (23.8; 88.2) |
| 1053 | Herzogtum Lauenburg | Rural district (with densification tendencies) | Schleswig-Holstein | 47288 | 22 | 46.5 | 46.3 | (30.4; 70.7) |
| 5112 | Duisburg, Stadt | Large urban city | Nordrhein-Westfalen | 66150 | 32 | 48.4 | 46.6 | (32.9; 65.9) |
| 6533 | Limburg-Weilburg | Urban district | Hessen | 38298 | 18 | 47.0 | 46.9 | (29.4; 74.6) |
| 5558 | Coesfeld | Urban district | Nordrhein-Westfalen | 48129 | 24 | 49.9 | 47.4 | (31.6; 71.0) |
| 5770 | Minden-Lübbecke | Urban district | Nordrhein-Westfalen | 37829 | 19 | 50.2 | 47.4 | (30.0; 74.8) |
| 5754 | Gütersloh | Urban district | Nordrhein-Westfalen | 42488 | 21 | 49.4 | 47.5 | (30.8; 73.1) |
| 5974 | Soest | Urban district | Nordrhein-Westfalen | 57064 | 28 | 49.1 | 47.6 | (32.8; 69.1) |
| 6435 | Main-Kinzig-Kreis | Urban district | Hessen | 81480 | 40 | 49.1 | 48.7 | (35.7; 66.5) |
| 5114 | Krefeld, Stadt | Large urban city | Nordrhein-Westfalen | 36760 | 18 | 49.0 | 49.3 | (31.1; 78.4) |
| 8118 | Ludwigsburg | Urban district | Baden-Württemberg | 77402 | 38 | 49.1 | 50.0 | (36.3; 68.8) |
| 5513 | Gelsenkirchen, Stadt | Large urban city | Nordrhein-Westfalen | 31326 | 16 | 51.1 | 50.3 | (30.8; 82.1) |

| District ID | Name of district | Urbanicity | Federal state | Database population, n | Clozapine users, n | Crude prevalence | Standardized prevalence | 95% CI |
|---|---|---|---|---|---|---|---|---|
| 7141 | Rhein-Lahn-Kreis | Urban district | Rheinland-Pfalz | 22913 | 11 | 48.0 | 50.4 | (27.7; 91.7) |
| 5915 | Hamm, Stadt | Large urban city | Nordrhein-Westfalen | 21870 | 11 | 50.3 | 50.6 | (27.9; 91.7) |
| 1058 | Rendsburg-Eckernförde | Rural district (with densification tendencies) | Schleswig-Holstein | 68843 | 35 | 50.8 | 51.2 | (36.6; 71.6) |
| 12060 | Barnim | Rural district (with densification tendencies) | Brandenburg | 35217 | 18 | 51.1 | 51.3 | (31.9; 82.6) |
| 9184 | München | Urban district | Bayern | 71312 | 34 | 47.7 | 51.6 | (36.7; 72.6) |
| 1060 | Segeberg | Rural district (with densification tendencies) | Schleswig-Holstein | 70401 | 37 | 52.6 | 52.6 | (38.1; 72.8) |
| 8115 | Böblingen | Urban district | Baden-Württemberg | 50013 | 26 | 52.0 | 52.9 | (36.0; 77.9) |
| 5382 | Rhein-Sieg-Kreis | Urban district | Nordrhein-Westfalen | 127708 | 67 | 52.5 | 53.1 | (41.8; 67.6) |
| 5962 | Märkischer Kreis | Urban district | Nordrhein-Westfalen | 63278 | 35 | 55.3 | 53.5 | (38.2; 74.9) |
| 9576 | Roth | Rural district (with densification tendencies) | Bayern | 23716 | 13 | 54.8 | 53.5 | (30.9; 92.6) |
| 8136 | Ostalbkreis | Urban district | Baden-Württemberg | 26663 | 14 | 52.5 | 53.9 | (31.8; 91.2) |
| 8116 | Esslingen | Urban district | Baden-Württemberg | 62692 | 34 | 54.2 | 54.0 | (38.5; 75.8) |
| 5158 | Mettmann | Urban district | Nordrhein-Westfalen | 75696 | 42 | 55.5 | 54.6 | (40.2; 74.1) |
| 5370 | Heinsberg | Urban district | Nordrhein-Westfalen | 37317 | 21 | 56.3 | 54.8 | (35.5; 84.4) |
| 3241 | Region Hannover | Urban district | Niedersachsen | 190741 | 107 | 56.1 | 55.5 | (45.9; 67.1) |
| 5116 | Mönchengladbach, St. | Large urban city | Nordrhein-Westfalen | 41803 | 23 | 55.0 | 55.5 | (36.9; 83.6) |
| 7131 | Ahrweiler | Rural district (with densification tendencies) | Rheinland-Pfalz | 21874 | 12 | 54.9 | 55.8 | (31.5; 98.7) |
| 5166 | Viersen | Urban district | Nordrhein-Westfalen | 51599 | 29 | 56.2 | 56.1 | (38.7; 81.2) |
| 5570 | Warendorf | Urban district | Nordrhein-Westfalen | 45446 | 26 | 57.2 | 56.2 | (38.2; 82.7) |
| 3355 | Lüneburg | Sparsely populated rural district | Niedersachsen | 34645 | 20 | 57.7 | 56.4 | (36.3; 87.7) |
| 6436 | Main-Taunus-Kreis | Urban district | Hessen | 62538 | 36 | 57.6 | 56.5 | (40.6; 78.5) |
| 5154 | Kleve | Urban district | Nordrhein-Westfalen | 48381 | 27 | 55.8 | 57.1 | (38.9; 83.7) |
| 3254 | Hildesheim | Urban district | Niedersachsen | 43116 | 24 | 55.7 | 57.3 | (38.3; 85.6) |
| 7137 | Mayen-Koblenz | Urban district | Rheinland-Pfalz | 31243 | 18 | 57.6 | 57.4 | (36.0; 91.5) |
| 3351 | Celle | Sparsely populated rural district | Niedersachsen | 24610 | 14 | 56.9 | 58.3 | (34.4; 98.8) |
| 6432 | Darmstadt-Dieburg | Urban district | Hessen | 65539 | 39 | 59.5 | 58.3 | (42.5; 80.0) |
| 1056 | Pinneberg | Urban district | Schleswig-Holstein | 91892 | 55 | 59.9 | 58.4 | (44.8; 76.2) |

| District ID | Name of district | Urbanicity | Federal state | Database population, n | Clozapine users, n | Crude prevalence | Standardized prevalence | 95% CI |
|---|---|---|---|---|---|---|---|---|
| 6440 | Wetteraukreis | Urban district | Hessen | 70582 | 42 | 59.5 | 58.6 | (43.2; 79.4) |
| 9187 | Rosenheim | Urban district | Bayern | 39713 | 22 | 55.4 | 58.6 | (38.4; 89.5) |
| 5162 | Rhein-Kreis Neuss | Urban district | Nordrhein-Westfalen | 81279 | 46 | 56.6 | 58.7 | (43.9; 78.5) |
| 1057 | Plön | Rural district (with densification tendencies) | Schleswig-Holstein | 31379 | 19 | 60.6 | 58.8 | (37.2; 92.9) |
| 8236 | Enzkreis | Urban district | Baden-Württemberg | 29729 | 17 | 57.2 | 58.9 | (36.5; 94.9) |
| 6611 | Kassel, Stadt | Large urban city | Hessen | 35621 | 20 | 56.1 | 59.0 | (37.9; 91.7) |
| 6412 | Frankfurt a.M., Stadt | Large urban city | Hessen | 170647 | 92 | 53.9 | 59.3 | (48.0; 73.3) |
| 9375 | Regensburg | Rural district (with densification tendencies) | Bayern | 23362 | 13 | 55.6 | 59.5 | (34.4; 102.9) |
| 6439 | Rheingau-Taunus-Kreis | Urban district | Hessen | 45394 | 27 | 59.5 | 60.3 | (41.2; 88.4) |
| 13003 | Rostock, Stadt | Large urban city | Mecklenburg-Vorpommern | 30592 | 20 | 65.4 | 60.8 | (39.0; 94.7) |
| 5315 | Köln, Stadt | Large urban city | Nordrhein-Westfalen | 251013 | 145 | 57.8 | 62.0 | (52.4; 73.3) |
| 2000 | Hamburg, Stadt | Large urban city | Hamburg | 562324 | 327 | 58.2 | 62.1 | (55.6; 69.4) |
| 5111 | Düsseldorf, Stadt | Large urban city | Nordrhein-Westfalen | 131220 | 77 | 58.7 | 62.1 | (49.4; 78.1) |
| 5911 | Bochum, Stadt | Large urban city | Nordrhein-Westfalen | 66198 | 41 | 61.9 | 64.3 | (47.2; 87.5) |
| 8117 | Göppingen | Urban district | Baden-Württemberg | 22517 | 15 | 66.6 | 64.6 | (38.9; 107.3) |
| 9761 | Augsburg, Stadt | Large urban city | Bayern | 42141 | 24 | 57.0 | 64.8 | (43.0; 97.7) |
| 7339 | Mainz-Bingen | Urban district | Rheinland-Pfalz | 46280 | 31 | 67.0 | 65.1 | (45.6; 92.8) |
| 13071 | Mecklenburg. Seenpl. | Sparsely populated rural district | Mecklenburg-Vorpommern | 39610 | 25 | 63.1 | 65.1 | (43.3; 97.9) |
| 6532 | Lahn-Dill-Kreis | Urban district | Hessen | 60131 | 39 | 64.9 | 65.4 | (47.7; 89.6) |
| 8111 | Stuttgart, Stadt | Large urban city | Baden-Württemberg | 97020 | 57 | 58.8 | 65.7 | (50.2; 86.1) |
| 5358 | Düren | Urban district | Nordrhein-Westfalen | 43559 | 28 | 64.3 | 65.8 | (45.4; 95.4) |
| 8119 | Rems-Murr-Kreis | Urban district | Baden-Württemberg | 48601 | 31 | 63.8 | 65.8 | (46.2; 93.7) |
| 9564 | Nürnberg, Stadt | Large urban city | Bayern | 77781 | 49 | 63.0 | 65.9 | (49.7; 87.5) |
| 7338 | Rhein-Pfalz-Kreis | Urban district | Rheinland-Pfalz | 27495 | 17 | 61.8 | 66.5 | (41.2; 107.4) |
| 5914 | Hagen, Stadt | Large urban city | Nordrhein-Westfalen | 30510 | 21 | 68.8 | 67.1 | (43.7; 102.9) |
| 3252 | Hameln-Pyrmont | Rural district (with densification tendencies) | Niedersachsen | 21568 | 14 | 64.9 | 68.8 | (40.4; 117.4) |
| 5562 | Recklinghausen | Urban district | Nordrhein-Westfalen | 86916 | 62 | 71.3 | 68.8 | (53.6; 88.3) |

| District ID | Name of district | Urbanicity | Federal state | Database population, n | Clozapine users, n | Crude prevalence | Standardized prevalence | 95% CI |
|---|---|---|---|---|---|---|---|---|
| 8417 | Zollernalbkreis | Urban district | Baden-Württemberg | 20971 | 16 | 76.3 | 69.3 | (42.1; 114.0) |
| 3257 | Schaumburg | Urban district | Niedersachsen | 20165 | 14 | 69.4 | 69.6 | (40.9; 118.5) |
| 14713 | Leipzig, Stadt | Large urban city | Sachsen | 65404 | 47 | 71.9 | 69.7 | (51.3; 94.7) |
| 6411 | Darmstadt, Stadt | Large urban city | Hessen | 43115 | 27 | 62.6 | 69.8 | (47.3; 103.0) |
| 6414 | Wiesbaden, Stadt | Large urban city | Hessen | 62565 | 44 | 70.3 | 70.3 | (52.2; 94.5) |
| 11000 | Berlin, Stadt | Large urban city | Berlin | 1013019 | 680 | 67.1 | 70.3 | (65.0; 75.9) |
| 8125 | Heilbronn | Urban district | Baden-Württemberg | 34025 | 24 | 70.5 | 70.7 | (47.4; 105.5) |
| 4011 | Bremen, Stadt | Large urban city | Bremen | 253163 | 171 | 67.5 | 71.0 | (61.1; 82.5) |
| 8216 | Rastatt | Urban district | Baden-Württemberg | 33898 | 25 | 73.8 | 71.9 | (48.4; 107.0) |
| 8416 | Tübingen | Urban district | Baden-Württemberg | 36361 | 24 | 66.0 | 72.4 | (48.2; 108.9) |
| 9679 | Würzburg | Urban district | Bayern | 30718 | 21 | 68.4 | 73.4 | (47.7; 113.1) |
| 8336 | Lörrach | Urban district | Baden-Württemberg | 33827 | 25 | 73.9 | 73.7 | (49.6; 109.5) |
| 5366 | Euskirchen | Urban district | Nordrhein-Westfalen | 28970 | 22 | 75.9 | 74.3 | (48.7; 113.5) |
| 3101 | Braunschweig, Stadt | Large urban city | Niedersachsen | 49713 | 35 | 70.4 | 75.1 | (53.6; 105.2) |
| 6431 | Bergstraße | Urban district | Hessen | 51152 | 40 | 78.2 | 75.5 | (55.3; 103.2) |
| 5774 | Paderborn | Urban district | Nordrhein-Westfalen | 44468 | 33 | 74.2 | 76.1 | (53.9; 107.3) |
| 6634 | Schwalm-Eder-Kreis | Rural district (with densification tendencies) | Hessen | 28149 | 21 | 74.6 | 76.3 | (49.5; 117.4) |
| 7340 | Südwestpfalz | Rural district (with densification tendencies) | Rheinland-Pfalz | 25303 | 21 | 83.0 | 76.6 | (49.4; 118.9) |
| 8315 | Breisg.-Hochschwarzw. | Urban district | Baden-Württemberg | 41330 | 32 | 77.4 | 76.8 | (54.1; 109.0) |
| 10044 | Saarlouis | Urban district | Saarland | 24081 | 19 | 78.9 | 77.1 | (49.0; 121.5) |
| 16053 | Jena, Stadt | Large urban city | Thüringen | 21132 | 16 | 75.7 | 77.5 | (46.5; 129.0) |
| 14612 | Dresden, Stadt | Large urban city | Sachsen | 64530 | 48 | 74.4 | 78.2 | (58.4; 104.6) |
| 5113 | Essen, Stadt | Large urban city | Nordrhein-Westfalen | 95383 | 74 | 77.6 | 78.7 | (62.5; 99.0) |
| 7331 | Alzey-Worms | Urban district | Rheinland-Pfalz | 23536 | 20 | 85.0 | 79.7 | (51.1; 124.3) |
| 8215 | Karlsruhe | Urban district | Baden-Württemberg | 74891 | 62 | 82.8 | 80.3 | (62.5; 103.2) |
| 5766 | Lippe | Urban district | Nordrhein-Westfalen | 44936 | 37 | 82.3 | 82.1 | (59.1; 114.0) |
| 9162 | München, Stadt | Large urban city | Bayern | 324505 | 224 | 69.0 | 82.9 | (72.2; 95.3) |
| 6413 | Offenbach a.M., Stadt | Large urban city | Hessen | 22727 | 18 | 79.2 | 83.0 | (51.9; 132.5) |
| 1002 | Kiel, Stadt | Large urban city | Schleswig-Holstein | 52208 | 38 | 72.8 | 83.5 | (60.2; 115.6) |

| District ID | Name of district | Urbanicity | Federal state | Database population, n | Clozapine users, n | Crude prevalence | Standardized prevalence | 95% CI |
|---|---|---|---|---|---|---|---|---|
| 5566 | Steinfurt | Urban district | Nordrhein-Westfalen | 83527 | 71 | 85.0 | 84.5 | (66.9; 106.8) |
| 7332 | Bad Dürkheim | Urban district | Rheinland-Pfalz | 25704 | 21 | 81.7 | 84.9 | (55.1; 130.8) |
| 5314 | Bonn, Stadt | Large urban city | Nordrhein-Westfalen | 81875 | 62 | 75.7 | 85.2 | (66.1; 109.8) |
| 5334 | Städteregion Aachen | Urban district | Nordrhein-Westfalen | 119637 | 93 | 77.7 | 85.9 | (69.9; 105.5) |
| 8316 | Emmendingen | Urban district | Baden-Württemberg | 23276 | 21 | 90.2 | 89.3 | (58.0; 137.4) |
| 3404 | Osnabrück, Stadt | Large urban city | Niedersachsen | 31621 | 28 | 88.5 | 90.0 | (62.0; 130.9) |
| 1059 | Schleswig-Flensburg | Sparsely populated rural district | Schleswig-Holstein | 42958 | 37 | 86.1 | 91.7 | (66.3; 126.9) |
| 16051 | Erfurt, Stadt | Large urban city | Thüringen | 22066 | 23 | 104.2 | 93.5 | (62.1; 140.8) |
| 1003 | Lübeck, Stadt | Large urban city | Schleswig-Holstein | 42693 | 40 | 93.7 | 94.0 | (68.8; 128.5) |
| 5124 | Wuppertal, Stadt | Large urban city | Nordrhein-Westfalen | 48435 | 43 | 88.8 | 94.3 | (69.8; 127.2) |
| 5711 | Bielefeld, Stadt | Large urban city | Nordrhein-Westfalen | 44174 | 40 | 90.6 | 94.3 | (69.1; 128.7) |
| 6534 | Marburg-Biedenkopf | Rural district (with densification tendencies) | Hessen | 53961 | 49 | 90.8 | 94.5 | (71.3; 125.3) |
| 6635 | Waldeck-Frankenberg | Sparsely populated rural district | Hessen | 27944 | 27 | 96.6 | 95.1 | (64.7; 139.6) |
| 5970 | Siegen-Wittgenstein | Urban district | Nordrhein-Westfalen | 40503 | 39 | 96.3 | 96.2 | (70.1; 132.0) |
| 8415 | Reutlingen | Urban district | Baden-Württemberg | 30199 | 31 | 102.7 | 97.1 | (68.2; 138.2) |
| 3357 | Rotenburg (Wümme) | Sparsely populated rural district | Niedersachsen | 28064 | 29 | 103.3 | 98.6 | (68.2; 142.5) |
| 6535 | Vogelsbergkreis | Sparsely populated rural district | Hessen | 20172 | 21 | 104.1 | 101.1 | (65.8; 155.4) |
| 1061 | Steinburg | Sparsely populated rural district | Schleswig-Holstein | 22436 | 24 | 107.0 | 104.0 | (69.4; 155.7) |
| 10041 | Reg.verb. Saarbrücken | Urban district | Saarland | 46283 | 49 | 105.9 | 105.0 | (79.2; 139.3) |
| 15002 | Halle (Saale), Stadt | Large urban city | Sachsen-Anhalt | 31376 | 34 | 108.4 | 107.0 | (75.9; 150.7) |
| 8317 | Ortenaukreis | Urban district | Baden-Württemberg | 61340 | 69 | 112.5 | 109.3 | (86.1; 138.6) |
| 3159 | Göttingen | Urban district | Niedersachsen | 58114 | 62 | 106.7 | 109.9 | (85.6; 141.3) |
| 5515 | Münster, Stadt | Large urban city | Nordrhein-Westfalen | 82690 | 84 | 101.6 | 114.7 | (92.1; 142.7) |
| 8212 | Karlsruhe, Stadt | Large urban city | Baden-Württemberg | 62287 | 67 | 107.6 | 116.9 | (91.7; 149.1) |
| 6531 | Gießen | Urban district | Hessen | 66878 | 78 | 116.6 | 117.5 | (93.9; 147.1) |
| 7315 | Mainz, Stadt | Large urban city | Rheinland-Pfalz | 53532 | 54 | 100.9 | 118.1 | (89.9; 155.1) |
| 8436 | Ravensburg | Urban district | Baden-Württemberg | 33547 | 41 | 122.2 | 118.5 | (87.0; 161.4) |
| 8311 | Freiburg i.Br., Stadt | Large urban city | Baden-Württemberg | 46690 | 49 | 104.9 | 120.3 | (90.1; 160.7) |
| 7337 | Südliche Weinstraße | Urban district | Rheinland-Pfalz | 24427 | 32 | 131.0 | 122.3 | (85.9; 174.1) |
| 8435 | Bodenseekreis | Urban district | Baden-Württemberg | 27976 | 34 | 121.5 | 125.6 | (89.5; 176.3) |

| District ID | Name of district | Urbanicity | Federal state | Database population, n | Clozapine users, n | Crude prevalence | Standardized prevalence | 95% CI |
|---|---|---|---|---|---|---|---|---|
| 8222 | Mannheim, Stadt | Large urban city | Baden-Württemberg | 50274 | 59 | 117.4 | 126.7 | (97.8; 164.3) |
| 8226 | Rhein-Neckar-Kreis | Urban district | Baden-Württemberg | 103495 | 149 | 144.0 | 139.3 | (118.5; 163.7) |
| 4012 | Bremerhaven, Stadt | Large urban city | Bremen | 56893 | 76 | 133.6 | 139.6 | (111.5; 174.8) |
| 9190 | Weilheim-Schongau | Rural district (with densification tendencies) | Bayern | 21292 | 29 | 136.2 | 144.8 | (100.6; 208.6) |
| 1055 | Ostholstein | Rural district (with densification tendencies) | Schleswig-Holstein | 32055 | 46 | 143.5 | 147.5 | (109.9; 198.0) |
| 5958 | Hochsauerlandkreis | Rural district (with densification tendencies) | Nordrhein-Westfalen | 43327 | 68 | 156.9 | 152.2 | (119.7; 193.4) |
| 9173 | Bad Tölz-Wolfratsh. | Sparsely populated rural district | Bayern | 20595 | 31 | 150.5 | 154.5 | (108.3; 220.5) |
| 9362 | Regensburg, Stadt | Large urban city | Bayern | 24360 | 33 | 135.5 | 162.1 | (113.8; 230.8) |
| 8335 | Konstanz | Urban district | Baden-Württemberg | 51560 | 83 | 161.0 | 164.8 | (132.7; 204.7) |
| 8221 | Heidelberg, Stadt | Large urban city | Baden-Württemberg | 34844 | 46 | 132.0 | 166.8 | (123.8; 224.7) |
| 7314 | Ludwigsh. a.R., Stadt | Large urban city | Rheinland-Pfalz | 22578 | 44 | 194.9 | 208.4 | (155.0; 280.2) |
| 9663 | Würzburg, Stadt | Large urban city | Bayern | 25762 | 45 | 174.7 | 209.0 | (154.5; 282.9) |

CI, confidence interval.

Estimates of the prevalence proportion of clozapine prescriptions are age- and sex-standardized to the population of Germany as of 31 December 2022.

The sample included 202 (out of 401) districts with ≥20,000 individuals in the database population. Based on the total population of these districts in official statistics, they represent approximately 73% of the overall German population in 2022.
